# Fish oil supplementation modifies the genetic potential for blood lipids

**DOI:** 10.1101/2023.09.22.23295987

**Authors:** Yitang Sun, Tryggvi McDonald, Abigail Baur, Huifang Xu, Naveen Brahman Bateman, Ye Shen, Changwei Li, Kaixiong Ye

**Affiliations:** Department of Genetics, Franklin College of Arts and Sciences, University of Georgia, Athens, GA, USA; Department of Epidemiology and Biostatistics, College of Public Health, University of Georgia, Athens, GA, USA; Department of Epidemiology, School of Public Health and Tropical Medicine, Tulane University, New Orleans, LA, USA; Institute of Bioinformatics, University of Georgia, Athens, GA, USA

## Abstract

**Background:** Dyslipidemia is a well-known risk factor for cardiovascular disease, which has been the leading cause of mortality worldwide. Although habitual intake of fish oil has been implicated in offering cardioprotective effects through triglyceride reduction, the interactions of fish oil with the genetic predisposition to dysregulated lipids remain elusive.

**Objectives:** We examined whether fish oil supplementation can modify the genetic potential for the circulating levels of four lipids, including total cholesterol, low-density lipoprotein cholesterol (LDL-C), high-density lipoprotein cholesterol (HDL-C), and triglycerides.

**Methods:** A total of 441,985 participants with complete genetic and phenotypic data from the UK Biobank were included in our study. Polygenic scores (PGS) were calculated in participants of diverse ancestries. Multivariable linear regression models were used to assess associations with adjustment for relevant risk factors.

**Results:** Fish oil supplementation mitigated genetic susceptibility to elevated levels of total cholesterol, LDL-C, and triglycerides, while amplifying genetic potential for increased HDL-C among 424,090 participants of European ancestry (*P*_interaction_ < 0.05). Consistent significant findings were obtained using PGS calculated based on multiple genome-wide association studies or alternative PGS methods. We also showed that fish oil significantly attenuated genetic predisposition to high triglycerides in African-ancestry participants.

**Conclusions:** Fish oil supplementation attenuated the genetic susceptibility to elevated blood levels of total cholesterol, LDL-C, and triglycerides, while accentuating genetic potential for higher HDL-C. These results suggest that fish oil may have a beneficial impact on modifying genome-wide genetic effects on elevated lipid levels in the general population.

## Introduction

Dyslipidemia is well-known to be a major risk factor for cardiovascular disease (CVD), the leading cause of death globally (1). Elevated levels of total cholesterol, low-density lipoprotein cholesterol (LDL-C), and triglyceride serve as major risk factors for increased risks of ischemic events, as shown in epidemiologic and Mendelian randomization studies (2–7). Conversely, a raised level of high-density lipoprotein cholesterol (HDL-C) may have protective roles against CVD, autoimmune disease, and cancers (8, 9). Numerous studies have found that fish oil supplementation, predominantly eicosapentaenoic acid (EPA) and docosahexaenoic acid (DHA), exerts cardioprotective effects through the reduction of triglyceride level (10–12). However, the roles of fish oil supplementation in modulating genetic susceptibility to elevated lipid levels remains insufficiently investigated.

In addition to environmental factors, studies have highlighted the essential role of genetic factors in dyslipidemia (13). Genetic effects have been estimated to explain more than 50% of the phenotypic variance in blood lipid levels (14). The differential effect of a genotype in individuals with different environmental exposures, known as gene-environment interaction, also plays a crucial role in determining outcomes (15, 16). A prior genome-wide interaction study identified four novel loci, whose associations with blood lipids were modified by fish oil supplementation (17). Expanding the previous single-variant analysis to a genome-scale, we hypothesized that the genome-wide genetic effects on blood lipid levels, captured by polygenic scores (PGS), could be modified by habitual fish oil supplementation.

In the current study, we aimed to explore the associations between fish oil supplementation status and genetic predispositions to high levels of total cholesterol, LDL-C, HDL-C, and triglyceride. Large-scale genome-wide association studies (GWAS) have identified numerous single nucleotide polymorphisms (SNPs) significantly associated with elevated lipid levels (18). Leveraging this information and employing novel PGS approaches, we aggregated individual lipid-associated SNPs into genome-scale PGS for estimating the genetic predispositions to high lipids among participants in UK Biobank (UKB). Then, we examined the interaction effects between fish oil supplementation and these PGS on the observed levels of blood lipids. While our primary analysis focused on participants of European (EUR) ancestry, we also investigated the interactions in African (AFR), Central/South Asian (CSA), and East Asian (EAS) populations.

## Methods

### Study population

This study was conducted using data from UKB (application number 48818), which is a prospective population-based study of over 0.5 million participants aged 37–73 years at recruitment from 2006 to 2010 across the United Kingdom (19). The UKB study received ethical approval from the National Health Service North West Centre for Research Ethics Committee, and all participants provided electronically signed consent before joining the study. Participants provided information on sociodemographic, lifestyle, and medical records via questionnaires at baseline. Blood samples for genotyping and biomarker measurements were also collected at recruitment. Of 502,369 individuals who attended baseline assessment, we excluded 6,705 individuals from the analysis due to missing fish oil supplementation data, mismatched information between phenotypic and genetic sex, sex chromosome aneuploidy, outliers for heterogeneity and missing genotype rate, and having a high degree of genetic kinship (ten or more third-degree relatives identified). Our analysis primarily focused on EUR ancestry participants, as they comprised the largest ancestral group in this resource. We also conducted analyses with participants of AFR, CSA, and EAS ancestries. Figure 1 presents a flowchart outlining the inclusion and exclusion of participants throughout our study.

**Figure 1.**
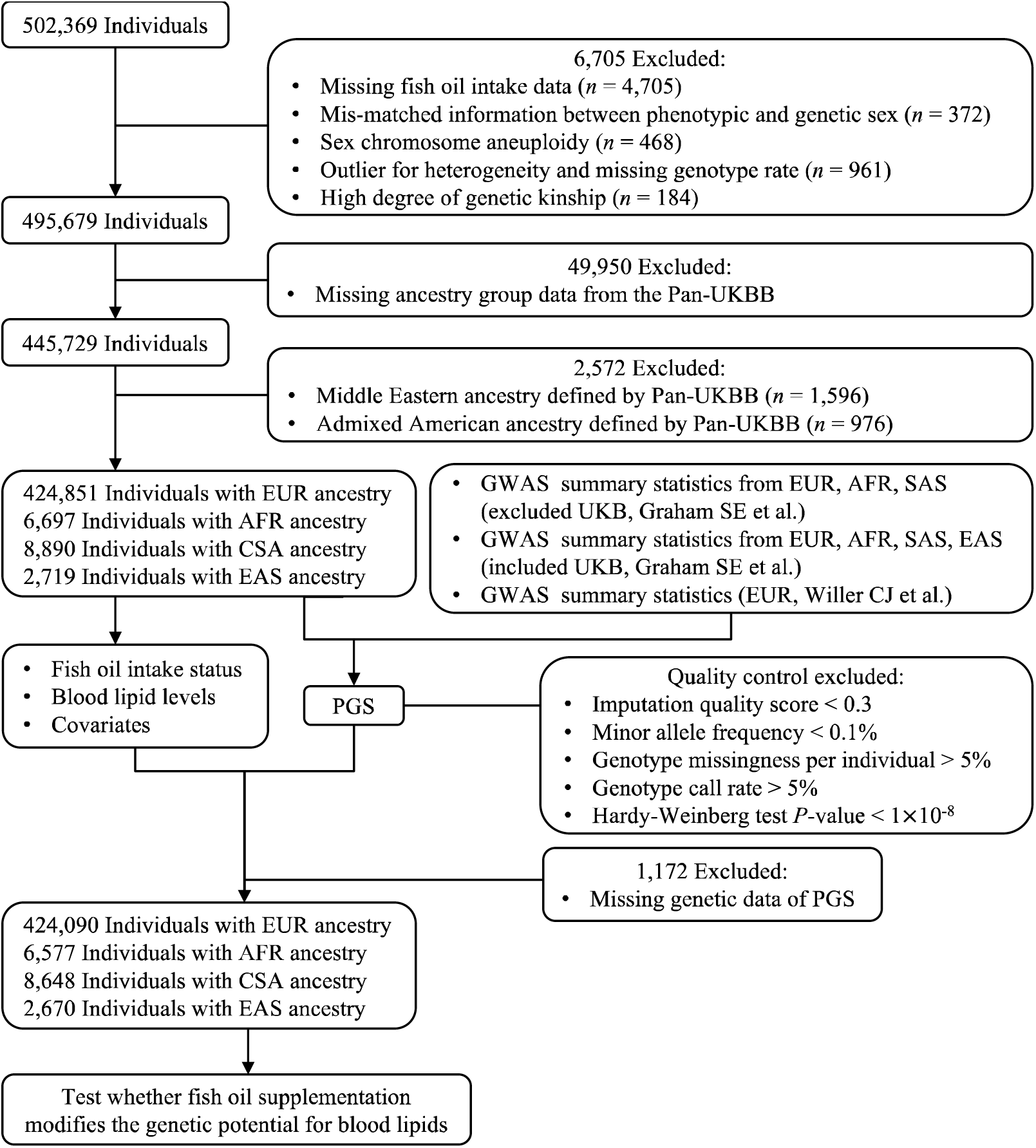
Flowchart of the study population selection. UKB, UK Biobank; Pan-UKBB, Pan-ancestry genetic analysis of the UK Biobank; EUR, European; AFR, African; CSA, Central/South Asian; EAS, East Asian; GWAS, genome-wide association study; PGS, polygenic scores.

### Assessment of fish oil supplementation status

The fish oil supplementation status of study participants was determined during their initial assessment center visit using a touchscreen questionnaire. Specifically, participants were asked about their regular consumption of certain supplements, with those reporting fish oil intake (including cod liver oil) classified as supplementing with fish oil.

### Assessment of blood lipids

Four baseline serum lipid parameters, including total cholesterol, LDL-C, HDL-C, and triglycerides, were ascertained using standard hematological methodologies. These lipids were directly quantified via a robust analytical platform, the Beckman Coulter AU5800 chemistry analyzer, to ensure precision and reproducibility.

### Polygenic score calculation

We computed PGS for participants of EUR, AFR, CSA, and EAS ancestries in the UKB, which measures the aggregated effects of genetic variants on total cholesterol, LDL-C, HDL-C, and triglycerides. In our primary discovery analysis, we used summary statistics from the largest and most recent GWAS to date, called the Graham SE *et al.* no UKB study, which excluded UKB and included up to 930,672 EUR, 92,555 AFR, and 34,135 CSA ancestry participants (20). In the replication analysis, we employed two additional GWAS summary statistics to calculate PGS. The first is the GWAS meta-analysis using UKB and other cohorts, which was released by Graham SE *et al.*, containing up to 1,320,016 EUR, 99,432 AFR, 40,963 CSA, and 146,492 EAS ancestry participants. The second is a GWAS that included up to 187,365 EUR individuals, as previously reported by Willer CJ *et al.* (21).

The PGS of UKB participants was calculated with the weighted method: weighted PGS = *β*_1_× SNP_1_ + *β*_2_ × SNP_2_ + … + *β*_n_ × SNP_n_, where SNP_n_ is the number of effect alleles of the n^th^ SNP, and the *β*-coefficient is the effect size of the corresponding allele, which was extracted from GWAS summary statistics. Quality control for genotypic data was based on the following criteria: (1) imputation quality score > 0.3, (2) minor allele frequency > 0.1%, (3) genotype missingness per individual < 5%, 4) genotype call rate < 5%, and (5) Hardy-Weinberg test *P*-value > 1 × 10^−8^. Independent significant SNPs were identified at a GWAS threshold of *P*-value < 5 × 10^−8^ with linkage disequilibrium (LD) clumping (*r*^2^ = 0.1, 250 kb). We also utilized the PGS *β*-coefficient previously generated by Weissbrod O *et al.* employing the methods of PolyPred, PolyPred+, and LD-pruning + *P*-value thresholding (P+T) (clump *r*^2^ = 0.5, 250 kb) (22). To facilitate the comparison of effects across four blood lipids, PGS was standardized to a mean of 0 and a standard deviation (SD) of 1, enabling interpretation per one SD change.

### Covariates

A series of sociodemographic, behavioral, and genetic covariates were included in our study. Age, sex, and body mass index (BMI) were obtained from baseline assessments at recruitment. The 22 baseline assessment center locations were situated throughout England, Scotland, and Wales. Socioeconomic status, as calculated by the Townsend deprivation index, was assigned to each participant corresponding to their home postcode at recruitment. Smoking and alcohol drinking status were defined as never, former, and current. Physical activity levels were categorized as low, moderate, and high following the International Physical Activity Questionnaire protocol. Statin use was ascertained through self-report during verbal interviews. Genetic data were generated through genotyping by either the UK Biobank Axiom Array or the UK BiLEVE Array. Participants were divided into ancestry groups based on the multi-ancestry analysis from the Pan-ancestry genetic analysis of the UK Biobank (Pan-UKBB). The top 20 ancestry principal components derived from the Pan-UKBB were employed as covariates.

### Statistical analysis

Multivariable linear regression models were conducted to assess the modifying impacts of fish oil supplementation on the genetic potential for blood lipids. Two groups of covariates were used to apply these models. In the first model, we adjusted for sex, age, age^2^, assessment centers, genotyping array, and the top 20 genetic principal components. The second model added additional covariates, including BMI, Townsend deprivation index, smoking status, alcohol status, physical activity, and statin use. We investigated the association between lipid PGS and lipid levels, stratifying by fish oil intake status among 424,090 participants of EUR ancestry to scrutinize the influence of fish oil supplementation on genetic predisposition for lipid levels. We further examined interactions between fish oil supplementation and PGS for four lipids on lipid levels by adding an interaction term between fish oil intake and lipid PGS and by incorporating both fish oil supplementation status and PGS as covariates in both models. Moreover, we conducted these interaction analyses using 6,577 AFR, 8,648 CSA, and 2,670 EAS ancestry participants. Serum levels of total cholesterol, LDL-C, HDL-C, and triglycerides were standardized, and their comparable effect sizes were expressed per one SD increase in the corresponding blood lipid. All analyses were conducted using R (version 4.2.1).

## Results

### Study cohorts

A total of 441,985 participants from the UKB with complete genetic and phenotypic data were included in this study, consisting mostly of individuals of EUR ancestry and a small proportion of other ancestries. We present the baseline characteristics of participants of EUR ancestry according to their fish oil intake status at baseline in Table 1. After quality control, 134,720 (31.8%) of study participants reported habitual use of fish oil supplements, while the remaining 289,370 participants were grouped as non-consumers of fish oil. The majority of fish oil users were women (56%), with a mean age of 59 years. Those in the fish oil intake group were more likely to be older, female, current alcohol drinkers, statin users, engaging in high physical activity, and having higher levels of total cholesterol, LDL-C, and HDL-C, but less likely to be current smokers or have an elevated BMI or triglyceride level. Among other ancestry groups, 2,289 (34.8%) AFR, 1,942 (22.5%) CSA, and 845 (31.6%) EAS participants took fish oil supplementation and displayed similar characteristics to the EUR participants (Supplementary Table 1).

**Table 1.**
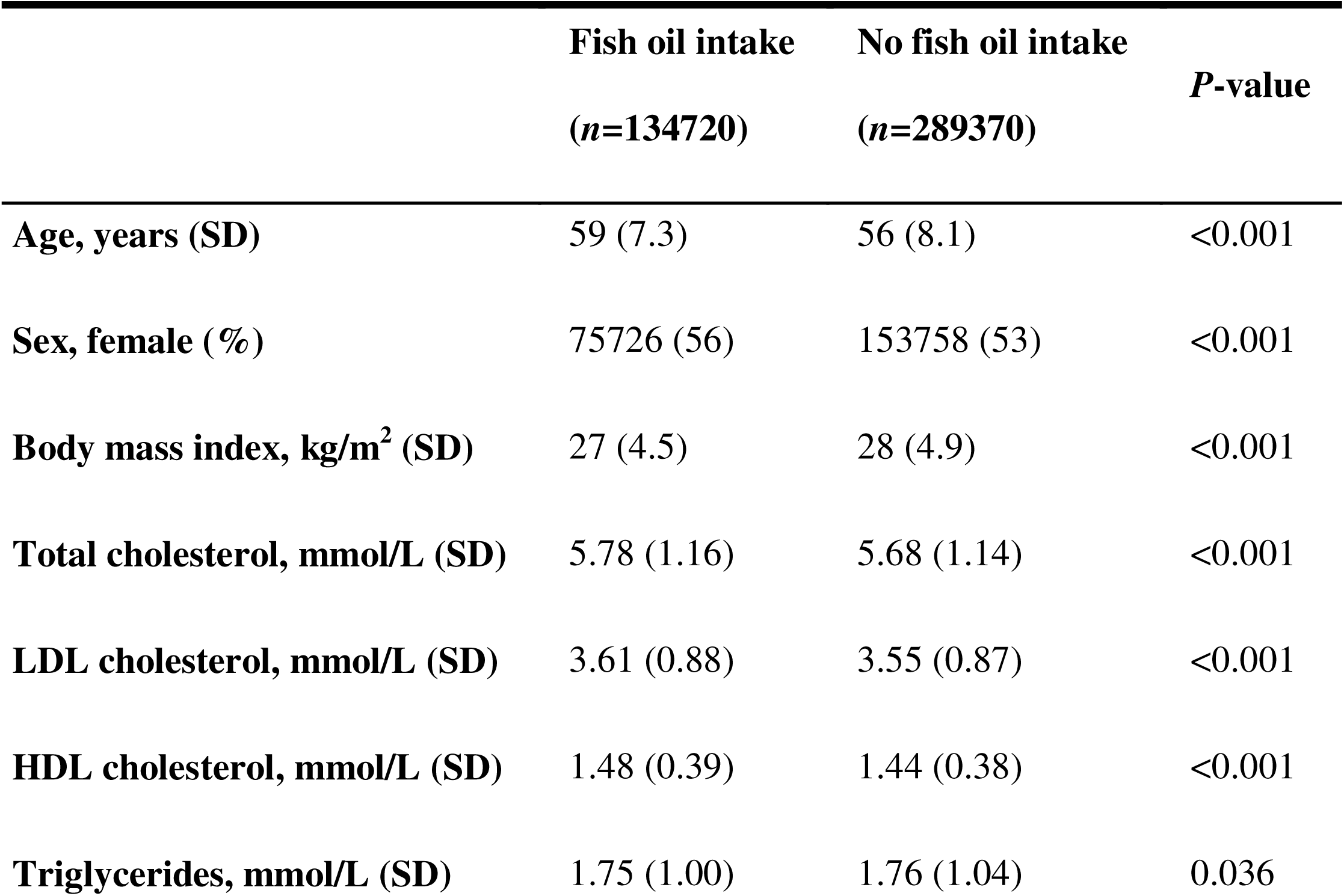
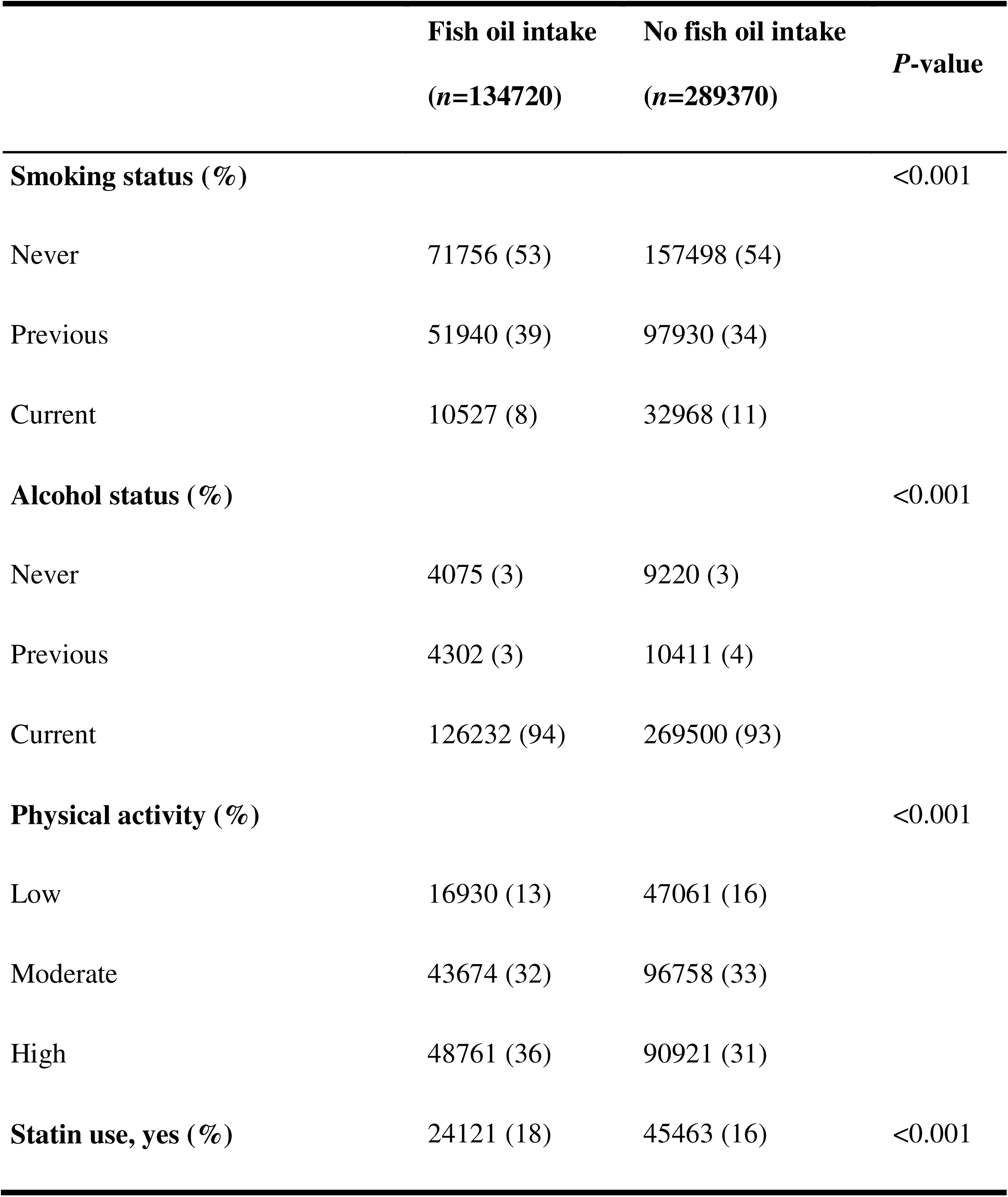
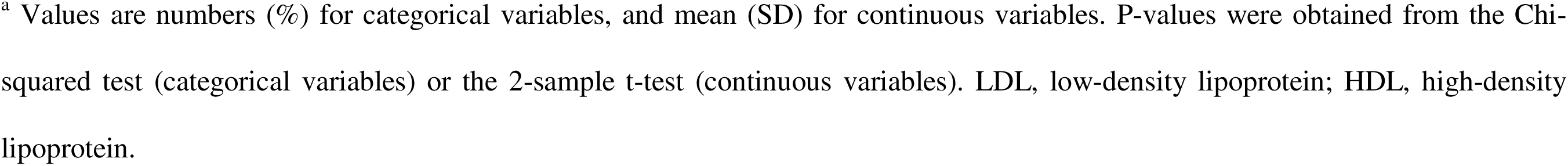
Baseline characteristics of participants of European ancestry in the UK Biobank^a^.

|  | **Fish oil intake**<br>**(*n*=134720)** | **No fish oil intake**<br>**(*n*=289370)** | ***P*-value** |
| --- | --- | --- | --- |
| **Age, years (SD)** | 59 (7.3) | 56 (8.1) | <0.001 |
| **Sex, female (%)** | 75726 (56) | 153758 (53) | <0.001 |
| **Body mass index, kg/m<sup>2</sup> (SD)** | 27 (4.5) | 28 (4.9) | <0.001 |
| **Total cholesterol, mmol/L (SD)** | 5.78 (1.16) | 5.68 (1.14) | <0.001 |
| **LDL cholesterol, mmol/L (SD)** | 3.61 (0.88) | 3.55 (0.87) | <0.001 |
| **HDL cholesterol, mmol/L (SD)** | 1.48 (0.39) | 1.44 (0.38) | <0.001 |
| **Triglycerides, mmol/L (SD)** | 1.75 (1.00) | 1.76 (1.04) | 0.036 |

|  | **Fish oil intake**<br>**(n=134720)** | **No fish oil intake**<br>**(n=289370)** | ***P*-value** |
| --- | --- | --- | --- |
| **Smoking status (%)** |  |  | <0.001 |
| Never | 71756 (53) | 157498 (54) |  |
| Previous | 51940 (39) | 97930 (34) |  |
| Current | 10527 (8) | 32968 (11) |  |
| **Alcohol status (%)** |  |  | <0.001 |
| Never | 4075 (3) | 9220 (3) |  |
| Previous | 4302 (3) | 10411 (4) |  |
| Current | 126232 (94) | 269500 (93) |  |
| **Physical activity (%)** |  |  | <0.001 |
| Low | 16930 (13) | 47061 (16) |  |
| Moderate | 43674 (32) | 96758 (33) |  |
| High | 48761 (36) | 90921 (31) |  |
| **Statin use, yes (%)** | 24121 (18) | 45463 (16) | <0.001 |

<sup>a</sup> Values are numbers (%) for categorical variables, and mean (SD) for continuous variables. P-values were obtained from the Chi-squared test (categorical variables) or the 2-sample t-test (continuous variables). LDL, low-density lipoprotein; HDL, high-density lipoprotein.

### Effect modification by fish oil supplementation on genetic predisposition to elevated lipids

In general, fish oil supplementation provided protection against genetic susceptibility to elevated levels of total cholesterol, LDL-C, and triglycerides, while it amplified genetic potential for higher HDL-C level. For the primary analysis using PGS (excluded UKB, Graham SE *et al.*), there was strong evidence that fish oil supplementation modified the effect of the genetic potential for four lipids in either the partially or fully adjusted model (*P*_interaction_ < 0.05) (Figure 2 and Table 2). When we generated PGS using two other GWAS summary statistics of Graham SE *et al.* (included UKB) and Willer CJ *et al.*, consistently significant results were observed for the four lipids (Supplementary Figure 1 and Supplementary Table 2). We also calculated PGS based on the *β*-coefficient previously generated by Weissbrod O *et al.* employing PolyPred, which is another PGS method employing functionally informed fine-mapping methods to obtain the posterior association coefficients. Utilizing PGS calculated via the PolyPred and P+T methods, we also discovered that fish oil supplementation significantly modified the genetic potential for elevated blood lipids (Supplementary Figure 2 and Supplementary Table 2).

**Figure 2.**
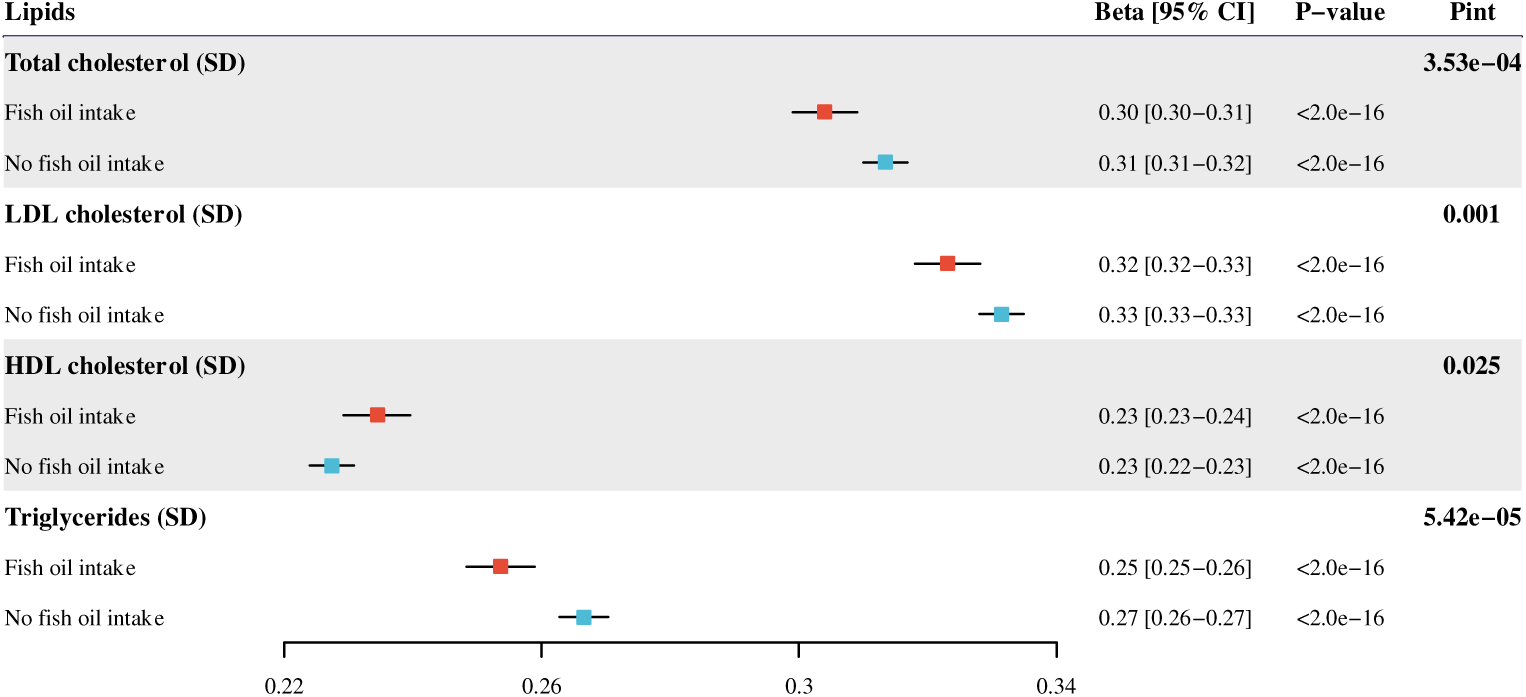
Interaction between fish oil and genetic predisposition to elevated blood lipids using GWAS summary statistics from Graham SE *et al.* (excluded UKB). Effect estimates and 95% confidence intervals are scaled to per 1 SD increase in PGS measured in SD units. UKB, UK Biobank; GWAS, genome-wide association study; PGS, polygenic scores; Pint, *P*-value for interaction.

**Table 2.**
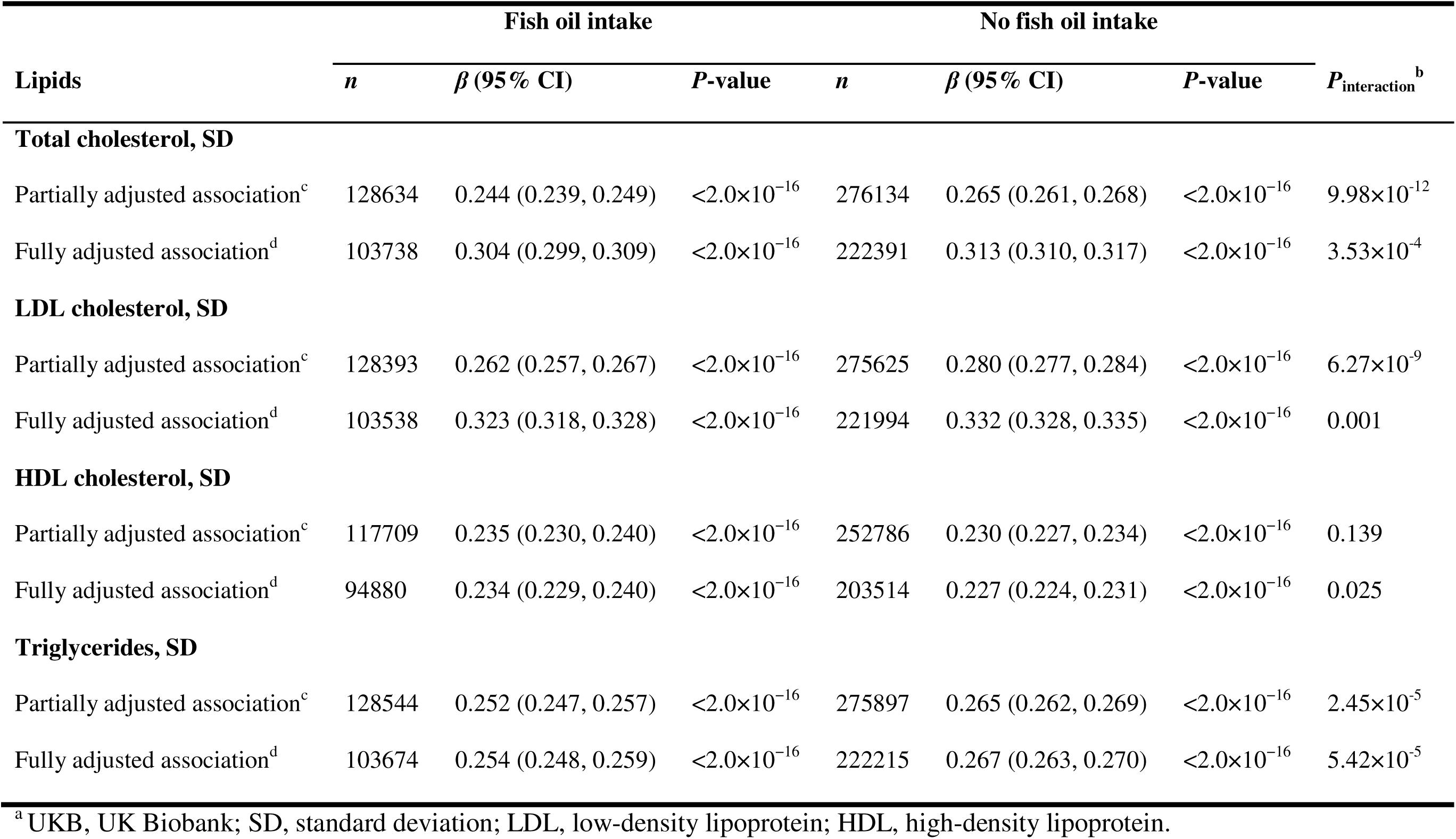
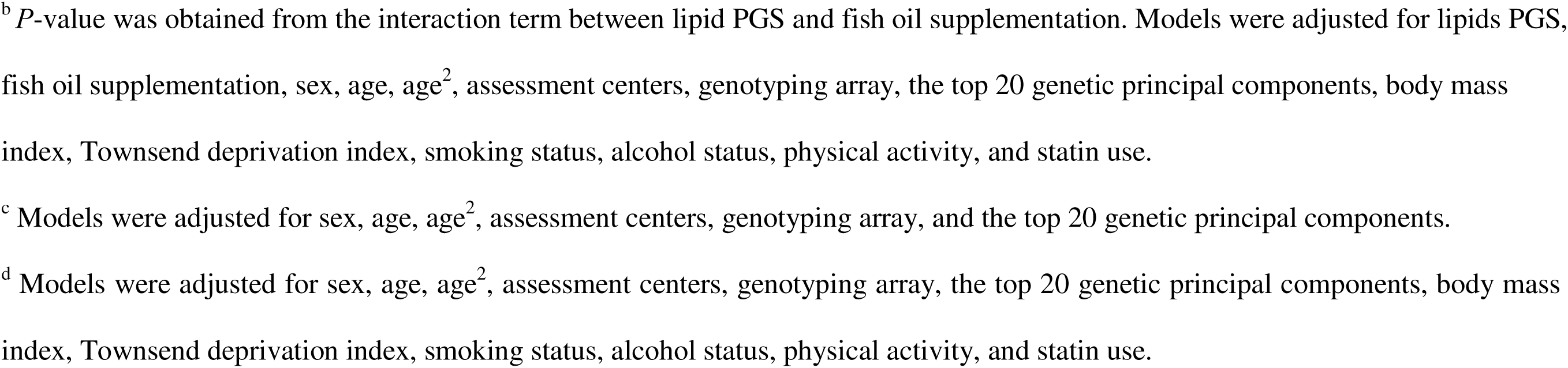
Association of lipid polygenic scores with observed lipid levels stratified by the fish oil intake status in UKB participants of European ancestry using GWAS summary statistics from Graham SE *et al.* (excluded UKB)^a^.

| Lipids | Fish oil intake | | | No fish oil intake | | | $P_{\text{interaction}}^{\text{b}}$ |
| --- | --- | --- | --- | --- | --- | --- | --- |
| | $n$ | $\beta$ (95% CI) | $P$-value | $n$ | $\beta$ (95% CI) | $P$-value | |
| **Total cholesterol, SD** |  |  |  |  |  |  |  |
| Partially adjusted association<sup>c</sup> | 128634 | 0.244 (0.239, 0.249) | $<2.0\times 10^{-16}$ | 276134 | 0.265 (0.261, 0.268) | $<2.0\times 10^{-16}$ | $9.98\times 10^{-12}$ |
| Fully adjusted association<sup>d</sup> | 103738 | 0.304 (0.299, 0.309) | $<2.0\times 10^{-16}$ | 222391 | 0.313 (0.310, 0.317) | $<2.0\times 10^{-16}$ | $3.53\times 10^{-4}$ |
| **LDL cholesterol, SD** |  |  |  |  |  |  |  |
| Partially adjusted association<sup>c</sup> | 128393 | 0.262 (0.257, 0.267) | $<2.0\times 10^{-16}$ | 275625 | 0.280 (0.277, 0.284) | $<2.0\times 10^{-16}$ | $6.27\times 10^{-9}$ |
| Fully adjusted association<sup>d</sup> | 103538 | 0.323 (0.318, 0.328) | $<2.0\times 10^{-16}$ | 221994 | 0.332 (0.328, 0.335) | $<2.0\times 10^{-16}$ | 0.001 |
| **HDL cholesterol, SD** |  |  |  |  |  |  |  |
| Partially adjusted association<sup>c</sup> | 117709 | 0.235 (0.230, 0.240) | $<2.0\times 10^{-16}$ | 252786 | 0.230 (0.227, 0.234) | $<2.0\times 10^{-16}$ | 0.139 |
| Fully adjusted association<sup>d</sup> | 94880 | 0.234 (0.229, 0.240) | $<2.0\times 10^{-16}$ | 203514 | 0.227 (0.224, 0.231) | $<2.0\times 10^{-16}$ | 0.025 |
| **Triglycerides, SD** |  |  |  |  |  |  |  |
| Partially adjusted association<sup>c</sup> | 128544 | 0.252 (0.247, 0.257) | $<2.0\times 10^{-16}$ | 275897 | 0.265 (0.262, 0.269) | $<2.0\times 10^{-16}$ | $2.45\times 10^{-5}$ |
| Fully adjusted association<sup>d</sup> | 103674 | 0.254 (0.248, 0.259) | $<2.0\times 10^{-16}$ | 222215 | 0.267 (0.263, 0.270) | $<2.0\times 10^{-16}$ | $5.42\times 10^{-5}$ |

<sup>a</sup> UKB, UK Biobank; SD, standard deviation; LDL, low-density lipoprotein; HDL, high-density lipoprotein.
<sup>b</sup> *P*-value was obtained from the interaction term between lipid PGS and fish oil supplementation. Models were adjusted for lipids PGS, fish oil supplementation, sex, age, age<sup>2</sup>, assessment centers, genotyping array, the top 20 genetic principal components, body mass index, Townsend deprivation index, smoking status, alcohol status, physical activity, and statin use.
<sup>c</sup> Models were adjusted for sex, age, age<sup>2</sup>, assessment centers, genotyping array, and the top 20 genetic principal components.
<sup>d</sup> Models were adjusted for sex, age, age<sup>2</sup>, assessment centers, genotyping array, the top 20 genetic principal components, body mass index, Townsend deprivation index, smoking status, alcohol status, physical activity, and statin use.

In the stratified analysis, fish oil supplementation significantly attenuated the genetic association with total cholesterol: each 1 SD increase in PGS was associated with a 0.304 SD increase (95% CI = 0.299 – 0.309) in total cholesterol among fish oil supplement users, compared to a 0.313 SD increase (95% CI = 0.310 – 0.317) among non-users (Figure 2 and Table 2). Similarly, for LDL-C, a 1 SD increment in PGS resulted in an increase of 0.323 SD (95% CI = 0.318 – 0.328) in LDL-C level among fish oil users, in comparison to 0.332 SD increase (95% CI = 0.328 – 0.335) for people without fish oil intake. Genetic susceptibility to high triglycerides was 0.254 (95% CI = 0.248 – 0.259) in the fish oil group and 0.267 (95% CI = 0.263 – 0.270) in the group not taking fish oil. Lastly, the impact of PGS was more pronounced in the fish oil group (0.234, 95% CI = 0.229 – 0.240) for HDL-C compared to participants without fish oil intake (0.227, 95% CI = 0.224 – 0.231). In summary, fish oil supplementation attenuates the genetic potential for elevated levels of total cholesterol, LDL-C, and triglycerides, while amplifying the genetic predisposition to a high level of HDL-C.

### Interaction effects in other ancestry groups

In participants of AFR ancestry from UKB, we found that fish oil supplementation was consistently associated with a diminished risk of genetic susceptibility to high triglycerides (Supplementary Table 3). When stratified by fish oil supplementation status, each SD increase in PGS correlated to a 0.099 SD (95% CI = 0.070 – 0.128) rise in triglyceride level among fish oil users, compared to a 0.150 SD (95% CI = 0.125 – 0.174) increase among those not taking fish oil. However, this modification effect of fish oil supplementation was not observed in participants of CSA and EAS ancestries (Supplementary Tables 4 and 5). Moreover, analyses involving participants from other ancestry groups did not indicate significant interaction effects on total cholesterol, LDL-C, and HDL-C. In these diverse populations, the influence of PGS on lipid levels remained statistically significant even after adjusting for fish oil supplementation and multiple covariates (Supplementary Table 6).

## Discussion

In this extensive prospective study involving 424,090 participants of EUR ancestry, our results indicated that fish oil supplementation could modify the genetic susceptibility to elevated levels of four lipids. Our findings suggested that fish oil supplementation attenuated genetic predispositions to high levels of total cholesterol, LDL-C, and triglycerides, while accentuating the genetic potential for increased HDL-C level. Analyzing 6,577 AFR, 8,648 CSA, and 2,670 EAS ancestry participants, no consistent significant interactions were found. However, for participants of AFR ancestry, our results indicated that fish oil supplementation mitigates the genetic risk on increased triglycerides. Collectively, our findings supported the hypothesis that genome-wide genetic effects on blood lipid levels, captured by PGS, could be modified by habitual fish oil supplementation.

Although substantial studies have highlighted the effects of fish oil supplements in improving the lipid profile in hyperlipidemic patients, their influence on genetic susceptibility to dyslipidemia remains unclear (2, 11, 23–26). A recent dose-response metalJanalysis of 90 randomized controlled trials with 72,598 participants demonstrated that DHAlJ+lJEPA supplementation (>2 g/d) reduced triglyceride level (27). Skulas-Ray AC *et al.* concluded that pharmacological doses of omega-3 fatty acids (>3 g/d total EPA+DHA) effectively decrease triglycerides and can be safely combined with other lipid-lowering agents, particularly statins (28). The Japan EPA Lipid Intervention Study, involving 14,981 hypercholesterolemia Japanese patients, found that 1.8 g/d EPA in combination with a statin led to lower triglycerides compared with statin therapy alone (29). In the current study of non-EUR ancestry groups, we also observed fish oil attenuates genetic susceptibility to elevated triglycerides among participants of AFR ancestry, but no significant interaction was detected among participants of CSA and EAS ancestries, which may be due to insufficient sample sizes. Large-scale studies covering more representative samples from diverse ancestries are needed to further validate our findings. Regarding genetic factors, some studies have investigated the interactions between fish oil and lipid-related genetic variants (17, 30–33). Our findings, leveraging cumulative effects of many genetic variants across the genome to assess individual genetic predisposition to dyslipidemia, affirmed the protective role of fish oil against genetic susceptibility to dyslipidemia.

The precise mechanisms underlying the observed interactions between fish oil supplementation and genetic predisposition to elevated lipid levels are not yet fully elucidated. These modifying effects could potentially be attributed to the multifaceted benefits derived from fish oil supplements. The putative mechanisms through which fish oil supplementation influences human lipoprotein metabolism include inhibition of very low-density lipoprotein (VLDL) triglyceride synthesis, decreased apoprotein B synthesis, enhancement of VLDL turnover via an increased fractional catabolic rate of VLDL, depression of LDL synthesis, and reduction of postprandial lipemia (34). Additionally, fish oil intake has been found to exert beneficial effects on several factors implicated in cardiovascular health, including inflammatory, oxidative, thrombotic, vascular, and arrhythmogenic parameters (11, 35–40). However, it is plausible that other mechanisms may also be involved in this modifying effect. Future functional studies are warranted to discern the underlying biological mechanisms.

To the best of our knowledge, this is the first study to evaluate the modifying effects of fish oil supplementation on genetic susceptibility to elevated lipid levels. The major strength of our study lies in the large number of participants of EUR ancestry, providing sufficient statistical power to detect significant interactions. Another strength of our study is the inclusion of other ancestry groups, where we confirmed that fish oil attenuated genetic susceptibility to high triglycerides in the AFR population. We used various GWAS summary statistics and PGS methodologies to calculate PGS and applied a fully adjusted model with a wealth of data on covariates, including socioeconomic characteristics, lifestyle factors, and statin use. Notably, the PGS for the primary analysis incorporated multiple SNPs from the largest and most recent GWAS across different cohorts, excluding any sample overlap with the UKB, thereby circumventing the potential issue of spurious candidate gene interactions (41). Population structure can lead to inflated estimates of PGS prediction (41). To address this, we adjusted all models for the top 20 genetic principal components to minimize potential bias. This comprehensive study design ensured the robustness and consistency of our findings.

Several potential limitations should also be considered. First, the self-reported fish oil intake is vulnerable to both measurement error and recall bias. Second, since fish oil supplementation status was assessed only at baseline, we were unable to evaluate the effects of longitudinal changes or extended duration of use. Third, our data lacked details on specific doses of fish oil supplementation, and future research is needed to discern the independent roles of DHA and EPA at specific doses. Fourth, UKB recruited people aged between 37 and 73 years at baseline, and our findings may not be generalizable to other age groups, such as children and adolescents (42). Fifth, though our study included multiple ancestries, caution is advised when extrapolating our findings to non-EUR populations, and future investigations with larger sample sizes are necessary. Lastly, given the observational design of our study, we cannot completely rule out reverse causation or residual confounding.

In conclusion, our findings reveal that fish oil supplementation attenuates genetic susceptibility to elevated blood levels of total cholesterol, LDL-C, and triglycerides, while accentuating genetic potential for higher HDL-C. Moreover, we detected fish oil modulates genetic predisposition to high triglycerides among participants of AFR ancestry. These findings suggest that fish oil could serve a beneficial role in modulating genetic susceptibility to elevated lipid levels in the general population. Further studies with larger sample sizes from diverse ancestry and accurate dose information of fish oil supplements are warranted to corroborate and expand our findings.

## Acknowledgments

The authors’ responsibilities were as follows—YS and KY: designed the study; YS: performed data analysis, prepared visualizations, and wrote the original draft of the manuscript; TM, AB, HX, and NBB: contributed to the data analysis; YS, YS, CL, and KY: provided statistical advice; YS and KY: interpreted the results; KY: critically revised the paper; all authors: read and approved the final manuscript and took responsibility for the integrity of the work as a whole.

## Disclosure

The authors declare that there is no conflict of interest.

## Funding

This work was funded by the University of Georgia Research Foundation and by the National Institute of General Medical Sciences of the National Institutes of Health under Award Number R35GM143060. The content is solely the responsibility of the authors and does not necessarily represent the official views of the National Institutes of Health.

## Data availability

Access to the UK Biobank resource is available via the application (http://www.ukbiobank.ac.uk). PGS coefficients generated by Graham SE *et al.* are available for public download at http://csg.sph.umich.edu/willer/public/glgc-lipids2021/results/ancestry_specific/. PGS coefficients generated by Willer CJ *et al.* are available for public download at http://ftp.ebi.ac.uk/pub/databases/gwas/summary_statistics/GCST002001-GCST003000/GCST002221/harmonised/. PGS coefficients generated by Weissbrod O *et al.* are available for public download at https://alkesgroup.broadinstitute.org/polypred_results.

## Supplementary Figures

**Supplementary Figure 1.** Interaction between fish oil and genetic predisposition to elevated blood lipids using GWAS summary statistics from Graham SE *et al.* (included UKB) and Willer CJ *et al.*.

Effect estimates and 95% confidence intervals are scaled to per 1 SD increase in PGS measured in SD units. UKB, UK Biobank; GWAS, genome-wide association study; PGS, polygenic scores; Pint, *P*-value for interaction.

**Supplementary Figure 2.** Interaction between fish oil and genetic predisposition to elevated blood lipids using the PGS *β*-coefficient generated by Weissbrod O *et al.*, including PolyPred and P+T.

Effect estimates and 95% confidence intervals are scaled to per 1 SD increase in PGS measured in SD units. PGS, polygenic scores; Pint, *P*-value for interaction.

## Supplementary Tables

**Supplementary Table 1.** Baseline characteristics of participants of African, Central/South Asian, and East Asian ancestries in the UK Biobank ^a^

^a^ Values are numbers (%) for categorical variables, and mean (SD) for continuous variables. LDL, low-density lipoprotein; HDL, high-density lipoprotein.

**Supplementary Table 2.** Association of lipid polygenic scores with observed lipid levels stratified by the fish oil intake status in participants of European ancestry^a^

^a^ UKB, UK Biobank; SD, standard deviation; LDL, low-density lipoprotein; HDL, high-density lipoprotein.

^b^ *P*-value was obtained from the interaction term between lipid PGS and fish oil supplementation. Models were adjusted for lipids PGS, fish oil supplementation, sex, age, age^2^, assessment centers, genotyping array, the top 20 genetic principal components, body mass index, Townsend deprivation index, smoking status, alcohol status, physical activity, and statin use.

^c^ Models were adjusted for sex, age, age^2^, assessment centers, genotyping array, and the top 20 genetic principal components.

^d^ Models were adjusted for sex, age, age^2^, assessment centers, genotyping array, the top 20 genetic principal components, body mass index, Townsend deprivation index, smoking status, alcohol status, physical activity, and statin use.

**Supplementary Table 3.** Association of lipid polygenic scores with observed lipid levels stratified by the fish oil intake status in participants of African ancestry^a^

^a^ UKB, UK Biobank; SD, standard deviation; LDL, low-density lipoprotein; HDL, high-density lipoprotein.

^b^ *P*-value was obtained from the interaction term between lipid PGS and fish oil supplementation. Models were adjusted for lipids PGS, fish oil supplementation, sex, age, age^2^, assessment centers, genotyping array, the top 20 genetic principal components, body mass index, Townsend deprivation index, smoking status, alcohol status, physical activity, and statin use.

^c^ Models were adjusted for sex, age, age^2^, assessment centers, genotyping array, and the top 20 genetic principal components.

^d^ Models were adjusted for sex, age, age^2^, assessment centers, genotyping array, the top 20 genetic principal components, body mass index, Townsend deprivation index, smoking status, alcohol status, physical activity, and statin use.

**Supplementary Table 4.** Association of lipid polygenic scores with observed lipid levels stratified by the fish oil intake status in participants of Central/South Asian ancestry^a^

^a^ UKB, UK Biobank; SD, standard deviation; LDL, low-density lipoprotein; HDL, high-density lipoprotein.

^b^ *P*-value was obtained from the interaction term between lipid PGS and fish oil supplementation. Models were adjusted for lipids PGS, fish oil supplementation, sex, age, age^2^, assessment centers, genotyping array, the top 20 genetic principal components, body mass index, Townsend deprivation index, smoking status, alcohol status, physical activity, and statin use.

^c^ Models were adjusted for sex, age, age^2^, assessment centers, genotyping array, and the top 20 genetic principal components.

^d^ Models were adjusted for sex, age, age^2^, assessment centers, genotyping array, the top 20 genetic principal components, body mass index, Townsend deprivation index, smoking status, alcohol status, physical activity, and statin use.

**Supplementary Table 5.** Association of lipid polygenic scores with observed lipid levels stratified by the fish oil intake status in participants of East Asian ancestry^a^

^a^ UKB, UK Biobank; SD, standard deviation; LDL, low-density lipoprotein; HDL, high-density lipoprotein.

^b^ *P*-value was obtained from the interaction term between lipid PGS and fish oil supplementation. Models were adjusted for lipids PGS, fish oil supplementation, sex, age, age^2^, assessment centers, genotyping array, the top 20 genetic principal components, body mass index, Townsend deprivation index, smoking status, alcohol status, physical activity, and statin use.

^c^ Models were adjusted for sex, age, age^2^, assessment centers, genotyping array, and the top 20 genetic principal components.

^d^ Models were adjusted for sex, age, age^2^, assessment centers, genotyping array, the top 20 genetic principal components, body mass index, Townsend deprivation index, smoking status, alcohol status, physical activity, and statin use.

**Supplementary Table 6.** Effects of fish oil supplementation and PGS on observed lipid levels in UKB participants of diverse ancestries^a^

^a^ UKB, UK Biobank; SD, standard deviation; LDL, low-density lipoprotein; HDL, high-density lipoprotein; EUR, European; AFR, African; CSA, Central/South Asian; EAS, East Asian.

^b^ Models were adjusted for sex, age, age^2^, assessment centers, genotyping array, and the top 20 genetic principal components.

^c^ Models were adjusted for sex, age, age^2^, assessment centers, genotyping array, the top 20 genetic principal components, body mass index, Townsend deprivation index, smoking status, alcohol status, physical activity, and statin use.

## Notes

### Competing Interest Statement

The authors have declared no competing interest.

### Author Declarations

The National Health Service North West Centre for Research Ethics Committee gave ethical approval for the UK Biobank Study. The data was accessed with an approved application (#48818).

