## Supplementary Tables for "Fish oil supplementation modifies the genetic potential for blood lipids"

Supplementary Material

**Supplementary Table 1.** Baseline characteristics of participants of African, Central/South Asian, and East Asian ancestries in the UK Biobank^a^

|  | **African** | | **Central/South Asian** | | **East Asian** | |
| --- | --- | --- | --- | --- | --- | --- |
|  | **Fish oil intake (N=2289)** | **No fish oil intake (N=4288)** | **Fish oil intake (N=1942)** | **No fish oil intake (N=6706)** | **Fish oil intake (N=845)** | **No fish oil intake (N=1825)** |
| **Age, years (SD)** | 54 (8.2) | 51 (7.9) | 55 (8.3) | 53 (8.4) | 54 (7.9) | 51 (7.7) |
| **Sex, female (%)** | 1495 (65) | 2401 (56) | 978 (50) | 3021 (45) | 608 (72) | 1161 (64) |
| **Body mass index, kg/m^2^ (SD)** | 30 (5.4) | 30 (5.5) | 27 (4.2) | 27 (4.5) | 24 (3.5) | 25 (3.9) |
| **Total cholesterol, mmol/L (SD)** | 5.27 (1.09) | 5.21 (1.09) | 5.34 (1.15) | 5.29 (1.12) | 5.75 (1.08) | 5.50 (1.03) |
| **LDL cholesterol, mmol/L (SD)** | 3.25 (0.83) | 3.25 (0.84) | 3.33 (0.87) | 3.34 (0.85) | 3.55 (0.82) | 3.40 (0.78) |
| **HDL cholesterol, mmol/L (SD)** | 1.49 (0.37) | 1.43 (0.36) | 1.30 (0.33) | 1.24 (0.31) | 1.53 (0.40) | 1.43 (0.36) |
| **Triglycerides, mmol/L (SD)** | 1.16 (0.65) | 1.19 (0.70) | 1.94 (1.19) | 1.97 (1.14) | 1.77 (1.11) | 1.78 (1.12) |
| **Smoking status (%)** |  |  |  |  |  |  |
| Never | 1641 (72) | 2950 (69) | 1487 (77) | 5231 (78) | 646 (76) | 1335 (73) |
| Previous | 374 (16) | 759 (18) | 288 (15) | 788 (12) | 136 (16) | 316 (17) |
| Current | 255 (11) | 544 (13) | 155 (8) | 614 (9) | 59 (7) | 165 (9) |
| **Alcohol status (%)** |  |  |  |  |  |  |
| Never | 311 (14) | 689 (16) | 564 (29) | 2941 (44) | 170 (20) | 386 (21) |
| Previous | 102 (4) | 243 (6) | 103 (5) | 366 (5) | 36 (4) | 86 (5) |
| Current | 1860 (81) | 3338 (78) | 1263 (65) | 3343 (50) | 637 (75) | 1350 (74) |
| **Physical activity (%)** |  |  |  |  |  |  |
| Low | 300 (13) | 754 (18) | 333 (17) | 1393 (21) | 113 (13) | 324 (18) |
| Moderate | 598 (26) | 1277 (30) | 595 (31) | 2062 (31) | 283 (33) | 589 (32) |
| High | 783 (34) | 1212 (28) | 638 (33) | 1600 (24) | 286 (34) | 500 (27) |
| **Statin use, yes (%)** | 349 (15) | 627 (15) | 575 (30) | 1660 (25) | 97 (11) | 183 (10) |

^a^ Values are numbers (%) for categorical variables, and mean (SD) for continuous variables. LDL, low-density lipoprotein; HDL, high-density lipoprotein.

**Supplementary Table 2.** Association of lipid polygenic scores with observed lipid levels stratified by the fish oil intake status in participants of European ancestry^a^

|  | **Fish oil intake** | | | **No fish oil intake** | | | |  |
| --- | --- | --- | --- | --- | --- | --- | --- | --- |
| **Lipids** | ***n*** | ***β* (95% CI)** | ***P*-value** | | ***n*** | ***β* (95% CI)** | ***P*-value** | ***P*_interaction_^b^** |
| ***Graham SE et al. (included UKB)*** |  |  |  | |  |  |  |  |
| **Total cholesterol, SD** |  |  |  | |  |  |  |  |
| Partially adjusted association^c^ | 128634 | 0.269 (0.264, 0.274) | <2.0×10^−16^ | | 276134 | 0.290 (0.287, 0.294) | <2.0×10^−16^ | 5.16×10^-12^ |
| Fully adjusted association^d^ | 103738 | 0.331 (0.326, 0.336) | <2.0×10^−16^ | | 222391 | 0.341 (0.338, 0.344) | <2.0×10^−16^ | 1.33×10^-4^ |
| **LDL cholesterol, SD** |  |  |  | |  |  |  |  |
| Partially adjusted association^c^ | 128393 | 0.279 (0.274, 0.284) | <2.0×10^−16^ | | 275625 | 0.299 (0.295, 0.302) | <2.0×10^−16^ | 3.61×10^-10^ |
| Fully adjusted association^d^ | 103538 | 0.344 (0.339, 0.349) | <2.0×10^−16^ | | 221994 | 0.352 (0.349, 0.356) | <2.0×10^−16^ | 0.001 |
| **HDL cholesterol, SD** |  |  |  | |  |  |  |  |
| Partially adjusted association^c^ | 117709 | 0.332 (0.327, 0.337) | <2.0×10^−16^ | | 252786 | 0.321 (0.317, 0.324) | <2.0×10^−16^ | 1.85×10^-4^ |
| Fully adjusted association^d^ | 94880 | 0.322 (0.317, 0.327) | <2.0×10^−16^ | | 203514 | 0.310 (0.307, 0.314) | <2.0×10^−16^ | 2.38×10^-5^ |
| **Triglycerides, SD** |  |  |  | |  |  |  |  |
| Partially adjusted association^c^ | 128544 | 0.273 (0.268, 0.278) | <2.0×10^−16^ | | 275897 | 0.289 (0.286, 0.293) | <2.0×10^−16^ | 1.35×10^-7^ |
| Fully adjusted association^d^ | 103674 | 0.273 (0.268, 0.278) | <2.0×10^−16^ | | 222215 | 0.289 (0.285, 0.292) | <2.0×10^−16^ | 1.67×10^-6^ |
| ***Willer CJ et al.*** |  |  |  | |  |  |  |  |
| **Total cholesterol, SD** |  |  |  | |  |  |  |  |
| Partially adjusted association^c^ | 128634 | 0.234 (0.229, 0.239) | <2.0×10^−16^ | | 276134 | 0.253 (0.250, 0.257) | <2.0×10^−16^ | 5.20×10^-10^ |
| Fully adjusted association^d^ | 103738 | 0.289 (0.284, 0.295) | <2.0×10^−16^ | | 222391 | 0.298 (0.295, 0.302) | <2.0×10^−16^ | 0.001 |
| **LDL cholesterol, SD** |  |  |  | |  |  |  |  |
| Partially adjusted association^c^ | 128393 | 0.242 (0.237, 0.247) | <2.0×10^−16^ | | 275625 | 0.261 (0.257, 0.264) | <2.0×10^−16^ | 6.85×10^-9^ |
| Fully adjusted association^d^ | 103538 | 0.297 (0.292, 0.302) | <2.0×10^−16^ | | 221994 | 0.307 (0.304, 0.311) | <2.0×10^−16^ | 3.79×10^-4^ |
| **HDL cholesterol, SD** |  |  |  | |  |  |  |  |
| Partially adjusted association^c^ | 117709 | 0.267 (0.262, 0.272) | <2.0×10^−16^ | | 252786 | 0.255 (0.252, 0.259) | <2.0×10^−16^ | 1.91×10^-4^ |
| Fully adjusted association^d^ | 94880 | 0.264 (0.259, 0.269) | <2.0×10^−16^ | | 203514 | 0.252 (0.249, 0.256) | <2.0×10^−16^ | 6.97×10^-5^ |
| **Triglycerides, SD** |  |  |  | |  |  |  |  |
| Partially adjusted association^c^ | 128544 | 0.200 (0.195, 0.205) | <2.0×10^−16^ | | 275897 | 0.209 (0.206, 0.213) | <2.0×10^−16^ | 0.002 |
| Fully adjusted association^d^ | 103674 | 0.201 (0.195, 0.206) | <2.0×10^−16^ | | 222215 | 0.212 (0.208, 0.216) | <2.0×10^−16^ | 4.78×10^-4^ |
| ***PolyPred*** |  |  |  | |  |  |  |  |
| **Total cholesterol, SD** |  |  |  | |  |  |  |  |
| Partially adjusted association^c^ | 128634 | 0.406 (0.401, 0.411) | <2.0×10^−16^ | | 276134 | 0.421 (0.418, 0.424) | <2.0×10^−16^ | 2.95×10^-7^ |
| Fully adjusted association^d^ | 103738 | 0.449 (0.445, 0.454) | <2.0×10^−16^ | | 222391 | 0.454 (0.450, 0.457) | <2.0×10^−16^ | 0.063 |
| **LDL cholesterol, SD** |  |  |  | |  |  |  |  |
| Partially adjusted association^c^ | 128393 | 0.409 (0.404, 0.414) | <2.0×10^−16^ | | 275625 | 0.425 (0.422, 0.429) | <2.0×10^−16^ | 1.12×10^-7^ |
| Fully adjusted association^d^ | 103538 | 0.461 (0.456, 0.466) | <2.0×10^−16^ | | 221994 | 0.466 (0.462, 0.469) | <2.0×10^−16^ | 0.039 |
| **HDL cholesterol, SD** |  |  |  | |  |  |  |  |
| Partially adjusted association^c^ | 117709 | 0.543 (0.539, 0.548) | <2.0×10^−16^ | | 252786 | 0.528 (0.525, 0.531) | <2.0×10^−16^ | 8.65×10^-10^ |
| Fully adjusted association^d^ | 94880 | 0.509 (0.505, 0.514) | <2.0×10^−16^ | | 203514 | 0.491 (0.488, 0.494) | <2.0×10^−16^ | 1.88×10^-14^ |
| **Triglycerides, SD** |  |  |  | |  |  |  |  |
| Partially adjusted association^c^ | 128544 | 0.448 (0.433, 0.442) | <2.0×10^−16^ | | 275897 | 0.455 (0.451, 0.458) | <2.0×10^−16^ | 2.34×10^-9^ |
| Fully adjusted association^d^ | 103674 | 0.425 (0.420, 0.430) | <2.0×10^−16^ | | 222215 | 0.439 (0.436, 0.443) | <2.0×10^−16^ | 8.84×10^-6^ |
| ***P+T*** |  |  |  | |  |  |  |  |
| **Total cholesterol, SD** |  |  |  | |  |  |  |  |
| Partially adjusted association^c^ | 128634 | 0.245 (0.239, 0.250) | <2.0×10^−16^ | | 276134 | 0.263 (0.260, 0.267) | <2.0×10^−16^ | 2.76×10^-9^ |
| Fully adjusted association^d^ | 103738 | 0.295 (0.290, 0.300) | <2.0×10^−16^ | | 222391 | 0.303 (0.299, 0.306) | <2.0×10^−16^ | 0.004 |
| **LDL cholesterol, SD** |  |  |  | |  |  |  |  |
| Partially adjusted association^c^ | 128393 | 0.256 (0.251, 0.261) | <2.0×10^−16^ | | 275625 | 0.273 (0.270, 0.277) | <2.0×10^−16^ | 6.76×10^-8^ |
| Fully adjusted association^d^ | 103538 | 0.308 (0.303, 0.313) | <2.0×10^−16^ | | 221994 | 0.316 (0.313, 0.320) | <2.0×10^−16^ | 0.002 |
| **HDL cholesterol, SD** |  |  |  | |  |  |  |  |
| Partially adjusted association^c^ | 117709 | 0.303 (0.298, 0.308) | <2.0×10^−16^ | | 252786 | 0.293 (0.290, 0.296) | <2.0×10^−16^ | 0.001 |
| Fully adjusted association^d^ | 94880 | 0.299 (0.294, 0.304) | <2.0×10^−16^ | | 203514 | 0.287 (0.284, 0.291) | <2.0×10^−16^ | 1.72×10^-4^ |
| **Triglycerides, SD** |  |  |  | |  |  |  |  |
| Partially adjusted association^c^ | 128544 | 0.250 (0.245, 0.255) | <2.0×10^−16^ | | 275897 | 0.262 (0.259, 0.266) | <2.0×10^−16^ | 1.03×10^-4^ |
| Fully adjusted association^d^ | 103674 | 0.252 (0.246, 0.257) | <2.0×10^−16^ | | 222215 | 0.263 (0.259, 0.267) | <2.0×10^−16^ | 3.63×10^-4^ |

^a^ UKB, UK Biobank; SD, standard deviation; LDL, low-density lipoprotein; HDL, high-density lipoprotein.

^b^ *P*-value was obtained from the interaction term between lipid PGS and fish oil supplementation. Models were adjusted for lipids PGS, fish oil supplementation, sex, age, age^2^, assessment centers, genotyping array, the top 20 genetic principal components, body mass index, Townsend deprivation index, smoking status, alcohol status, physical activity, and statin use.

^c^ Models were adjusted for sex, age, age^2^, assessment centers, genotyping array, and the top 20 genetic principal components.

^d^ Models were adjusted for sex, age, age^2^, assessment centers, genotyping array, the top 20 genetic principal components, body mass index, Townsend deprivation index, smoking status, alcohol status, physical activity, and statin use.

**Supplementary Table 3.** Association of lipid polygenic scores with observed lipid levels stratified by the fish oil intake status in participants of African ancestry^a^

|  | **Fish oil intake** | | | **No fish oil intake** | | |  |
| --- | --- | --- | --- | --- | --- | --- | --- |
| **Lipids** | ***n*** | ***β* (95% CI)** | ***P*-value** | ***n*** | ***β* (95% CI)** | ***P*-value** | ***P*_interaction_^b^** |
| ***Graham SE et al. (excluded UKB)*** |  |  |  |  |  |  |  |
| **Total cholesterol, SD** |  |  |  |  |  |  |  |
| Partially adjusted association^c^ | 2145 | 0.315 (0.276, 0.355) | 1.83×10^-52^ | 4010 | 0.311 (0.282, 0.340) | 2.11×10^-93^ | 0.918 |
| Fully adjusted association^d^ | 1526 | 0.377 (0.333, 0.422) | 9.06×10^-57^ | 2978 | 0.341 (0.309, 0.374) | 1.78×10^-88^ | 0.496 |
| **LDL cholesterol, SD** |  |  |  |  |  |  |  |
| Partially adjusted association^c^ | 2140 | 0.345 (0.305, 0.385) | 1.86×10^-60^ | 4003 | 0.335 (0.306, 0.364) | 1.10×10^-106^ | 0.983 |
| Fully adjusted association^d^ | 1521 | 0.403 (0.357, 0.448) | 1.44×10^-61^ | 2973 | 0.376 (0.343, 0.408) | 5.40×10^-106^ | 0.721 |
| **HDL cholesterol, SD** |  |  |  |  |  |  |  |
| Partially adjusted association^c^ | 1997 | 0.234 (0.194, 0.275) | 5.55×10^-29^ | 3703 | 0.247 (0.218, 0.275) | 1.71×10^-63^ | 0.569 |
| Fully adjusted association^d^ | 1417 | 0.224 (0.178, 0.269) | 2.02×10^-21^ | 2748 | 0.238 (0.208, 0.267) | 3.45×10^-53^ | 0.676 |
| **Triglycerides, SD** |  |  |  |  |  |  |  |
| Partially adjusted association^c^ | 2144 | 0.089 (0.064, 0.114) | 3.32×10^-12^ | 4010 | 0.137 (0.116, 0.158) | 1.36×10^-36^ | 0.005 |
| Fully adjusted association^d^ | 1525 | 0.099 (0.070, 0.128) | 2.94×10^-11^ | 2978 | 0.150 (0.125, 0.174) | 4.90×10^-32^ | 0.007 |
| ***Graham SE et al. (included UKB)*** |  |  |  |  |  |  |  |
| **Total cholesterol, SD** |  |  |  |  |  |  |  |
| Partially adjusted association^c^ | 2145 | 0.339 (0.300, 0.379) | 5.51×10^-59^ | 4010 | 0.330 (0.301, 0.359) | 2.10×10^-104^ | 0.959 |
| Fully adjusted association^d^ | 1526 | 0.402 (0.357, 0.447) | 7.41×10^-63^ | 2978 | 0.364 (0.331, 0.396) | 3.78×10^-99^ | 0.479 |
| **LDL cholesterol, SD** |  |  |  |  |  |  |  |
| Partially adjusted association^c^ | 2140 | 0.359 (0.318, 0.399) | 2.94×10^-64^ | 4003 | 0.348 (0.319, 0.378) | 3.59×10^-113^ | 0.954 |
| Fully adjusted association^d^ | 1521 | 0.414 (0.369, 0.460) | 1.03×10^-64^ | 2973 | 0.391 (0.358, 0.423) | 1.74×10^-113^ | 0.890 |
| **HDL cholesterol, SD** |  |  |  |  |  |  |  |
| Partially adjusted association^c^ | 1997 | 0.254 (0.213, 0.294) | 1.01×10^-33^ | 3703 | 0.264 (0.237, 0.292) | 2.00×10^-73^ | 0.615 |
| Fully adjusted association^d^ | 1417 | 0.245 (0.200, 0.290) | 1.57×10^-25^ | 2748 | 0.252 (0.223, 0.281) | 3.33×10^-60^ | 0.872 |
| **Triglycerides, SD** |  |  |  |  |  |  |  |
| Partially adjusted association^c^ | 2144 | 0.105 (0.080, 0.130) | 3.86×10^-16^ | 4010 | 0.150 (0.129, 0.171) | 3.44×10^-44^ | 0.008 |
| Fully adjusted association^d^ | 1525 | 0.111 (0.082, 0.141) | 1.07×10^-13^ | 2978 | 0.159 (0.134, 0.183) | 5.15×10^-36^ | 0.011 |
| ***PolyPred+*** |  |  |  |  |  |  |  |
| **Total cholesterol, SD** |  |  |  |  |  |  |  |
| Partially adjusted association^c^ | 2145 | 0.293 (0.255, 0.331) | 5.63×10^-49^ | 4010 | 0.249 (0.220, 0.278) | 1.36×10^-62^ | 0.108 |
| Fully adjusted association^d^ | 1526 | 0.312 (0.269, 0.355) | 5.64×10^-43^ | 2978 | 0.275 (0.243, 0.307) | 4.11×10^-61^ | 0.243 |
| **LDL cholesterol, SD** |  |  |  |  |  |  |  |
| Partially adjusted association^c^ | 2140 | 0.337 (0.299, 0.374) | 6.98×10^-65^ | 4003 | 0.297 (0.268, 0.326) | 1.16×10^-85^ | 0.151 |
| Fully adjusted association^d^ | 1521 | 0.356 (0.314, 0.399) | 4.61×10^-56^ | 2973 | 0.329 (0.297, 0.360) | 3.54×10^-85^ | 0.357 |
| **HDL cholesterol, SD** |  |  |  |  |  |  |  |
| Partially adjusted association^c^ | 1997 | 0.268 (0.228, 0.308) | 7.04×10^-38^ | 3703 | 0.238 (0.211, 0.266) | 6.38×10^-61^ | 0.245 |
| Fully adjusted association^d^ | 1417 | 0.269 (0.224, 0.314) | 1.21×10^-30^ | 2748 | 0.231 (0.201, 0.260) | 2.08×10^-50^ | 0.157 |
| **Triglycerides, SD** |  |  |  |  |  |  |  |
| Partially adjusted association^c^ | 2144 | 0.082 (0.055, 0.108) | 2.05×10^-9^ | 4010 | 0.113 (0.092, 0.134) | 1.52×10^-26^ | 0.071 |
| Fully adjusted association^d^ | 1525 | 0.083 (0.051, 0.115) | 3.08×10^-7^ | 2978 | 0.114 (0.090, 0.138) | 3.21×10^-20^ | 0.122 |

^a^ UKB, UK Biobank; SD, standard deviation; LDL, low-density lipoprotein; HDL, high-density lipoprotein.

^b^ *P*-value was obtained from the interaction term between lipid PGS and fish oil supplementation. Models were adjusted for lipids PGS, fish oil supplementation, sex, age, age^2^, assessment centers, genotyping array, the top 20 genetic principal components, body mass index, Townsend deprivation index, smoking status, alcohol status, physical activity, and statin use.

^c^ Models were adjusted for sex, age, age^2^, assessment centers, genotyping array, and the top 20 genetic principal components.

^d^ Models were adjusted for sex, age, age^2^, assessment centers, genotyping array, the top 20 genetic principal components, body mass index, Townsend deprivation index, smoking status, alcohol status, physical activity, and statin use.

**Supplementary Table 4.** Association of lipid polygenic scores with observed lipid levels stratified by the fish oil intake status in participants of Central/South Asian ancestry^a^

|  | **Fish oil intake** | | | **No fish oil intake** | | |  |
| --- | --- | --- | --- | --- | --- | --- | --- |
| **Lipids** | ***n*** | ***β* (95% CI)** | ***P*-value** | ***n*** | ***β* (95% CI)** | ***P*-value** | ***P*_interaction_^b^** |
| ***Graham SE et al. (excluded UKB)*** |  |  |  |  |  |  |  |
| **Total cholesterol, SD** |  |  |  |  |  |  |  |
| Partially adjusted association^c^ | 1841 | 0.157 (0.111, 0.203) | 2.11×10^-11^ | 6373 | 0.156 (0.132, 0.179) | 8.72×10^-39^ | 0.883 |
| Fully adjusted association^d^ | 1445 | 0.208 (0.162, 0.254) | 1.58×10^-18^ | 4623 | 0.206 (0.182, 0.231) | 5.59×10^-59^ | 0.835 |
| **LDL cholesterol, SD** |  |  |  |  |  |  |  |
| Partially adjusted association^c^ | 1835 | 0.162 (0.117, 0.206) | 1.83×10^-12^ | 6363 | 0.152 (0.129, 0.176) | 5.58×10^-37^ | 0.793 |
| Fully adjusted association^d^ | 1441 | 0.204 (0.160, 0.248) | 3.47×10^-19^ | 4615 | 0.203 (0.179, 0.228) | 2.97×10^-58^ | 0.956 |
| **HDL cholesterol, SD** |  |  |  |  |  |  |  |
| Partially adjusted association^c^ | 1685 | 0.210 (0.172, 0.248) | 9.30×10^-27^ | 5807 | 0.185 (0.167, 0.204) | 1.21×10^-81^ | 0.122 |
| Fully adjusted association^d^ | 1330 | 0.210 (0.169, 0.252) | 1.29×10^-22^ | 4214 | 0.183 (0.162, 0.204) | 3.84×10^-64^ | 0.127 |
| **Triglycerides, SD** |  |  |  |  |  |  |  |
| Partially adjusted association^c^ | 1839 | 0.318 (0.265, 0.371) | 9.45×10^-31^ | 6370 | 0.265 (0.239, 0.291) | 4.55×10^-86^ | 0.070 |
| Fully adjusted association^d^ | 1444 | 0.303 (0.244, 0.362) | 5.46×10^-23^ | 4621 | 0.254 (0.224, 0.285) | 4.06×10^-59^ | 0.094 |
| ***Graham SE et al. (included UKB)*** |  |  |  |  |  |  |  |
| **Total cholesterol, SD** |  |  |  |  |  |  |  |
| Partially adjusted association^c^ | 1841 | 0.156 (0.111, 0.201) | 1.42×10^-11^ | 6373 | 0.164 (0.140, 0.187) | 1.01×10^-42^ | 0.846 |
| Fully adjusted association^d^ | 1445 | 0.206 (0.161, 0.252) | 9.03×10^-19^ | 4623 | 0.217 (0.192, 0.241) | 6.11×10^-65^ | 0.833 |
| **LDL cholesterol, SD** |  |  |  |  |  |  |  |
| Partially adjusted association^c^ | 1835 | 0.169 (0.124, 0.214) | 2.27×10^-13^ | 6363 | 0.162 (0.139, 0.185) | 7.25×10^-42^ | 0.913 |
| Fully adjusted association^d^ | 1441 | 0.223 (0.179, 0.267) | 1.81×10^-22^ | 4615 | 0.213 (0.189, 0.237) | 2.13×10^-64^ | 0.810 |
| **HDL cholesterol, SD** |  |  |  |  |  |  |  |
| Partially adjusted association^c^ | 1685 | 0.228 (0.191, 0.266) | 2.21×10^-31^ | 5807 | 0.199 (0.181, 0.218) | 4.21×10^-94^ | 0.072 |
| Fully adjusted association^d^ | 1330 | 0.234 (0.193, 0.275) | 9.65×10^-28^ | 4214 | 0.200 (0.179, 0.220) | 1.82×10^-76^ | 0.056 |
| **Triglycerides, SD** |  |  |  |  |  |  |  |
| Partially adjusted association^c^ | 1839 | 0.323 (0.271, 0.376) | 3.22×10^-32^ | 6370 | 0.276 (0.250, 0.302) | 1.80×10^-93^ | 0.104 |
| Fully adjusted association^d^ | 1444 | 0.304 (0.246, 0.362) | 5.24×10^-24^ | 4621 | 0.269 (0.238, 0.299) | 3.69×10^-65^ | 0.204 |
| ***PolyPred+*** |  |  |  |  |  |  |  |
| **Total cholesterol, SD** |  |  |  |  |  |  |  |
| Partially adjusted association^c^ | 1841 | 0.200 (0.156, 0.244) | 9.51×10^-19^ | 6373 | 0.207 (0.184, 0.230) | 2.03×10^-67^ | 0.784 |
| Fully adjusted association^d^ | 1445 | 0.271 (0.228, 0.314) | 2.44×10^-33^ | 4623 | 0.279 (0.255, 0.303) | 2.79×10^-108^ | 0.904 |
| **LDL cholesterol, SD** |  |  |  |  |  |  |  |
| Partially adjusted association^c^ | 1835 | 0.216 (0.172, 0.259) | 8.01×10^-22^ | 6363 | 0.206 (0.182, 0.229) | 1.37×10^-65^ | 0.773 |
| Fully adjusted association^d^ | 1441 | 0.288 (0.246, 0.330) | 2.89×10^-39^ | 4615 | 0.281 (0.257, 0.306) | 2.77×10^-110^ | 0.807 |
| **HDL cholesterol, SD** |  |  |  |  |  |  |  |
| Partially adjusted association^c^ | 1685 | 0.307 (0.272, 0.343) | 1.66×10^-59^ | 5807 | 0.289 (0.271, 0.307) | 3.04×10^-202^ | 0.289 |
| Fully adjusted association^d^ | 1330 | 0.327 (0.289, 0.365) | 2.62×10^-57^ | 4214 | 0.290 (0.270, 0.310) | 9.21×10^-165^ | 0.074 |
| **Triglycerides, SD** |  |  |  |  |  |  |  |
| Partially adjusted association^c^ | 1839 | 0.285 (0.234, 0.336) | 5.76×10^-27^ | 6370 | 0.280 (0.254, 0.306) | 1.54×10^-94^ | 0.745 |
| Fully adjusted association^d^ | 1444 | 0.271 (0.214, 0.328) | 4.56×10^-20^ | 4621 | 0.271 (0.240, 0.301) | 3.74×10^-65^ | 0.750 |

^a^ UKB, UK Biobank; SD, standard deviation; LDL, low-density lipoprotein; HDL, high-density lipoprotein.

^b^ *P*-value was obtained from the interaction term between lipid PGS and fish oil supplementation. Models were adjusted for lipids PGS, fish oil supplementation, sex, age, age^2^, assessment centers, genotyping array, the top 20 genetic principal components, body mass index, Townsend deprivation index, smoking status, alcohol status, physical activity, and statin use.

^c^ Models were adjusted for sex, age, age^2^, assessment centers, genotyping array, and the top 20 genetic principal components.

^d^ Models were adjusted for sex, age, age^2^, assessment centers, genotyping array, the top 20 genetic principal components, body mass index, Townsend deprivation index, smoking status, alcohol status, physical activity, and statin use.

**Supplementary Table 5.** Association of lipid polygenic scores with observed lipid levels stratified by the fish oil intake status in participants of East Asian ancestry^a^

|  | **Fish oil intake** | | | **No fish oil intake** | | |  |
| --- | --- | --- | --- | --- | --- | --- | --- |
| **Lipids** | ***n*** | ***β* (95% CI)** | ***P*-value** | ***n*** | ***β* (95% CI)** | ***P*-value** | ***P*_interaction_^b^** |
| ***Graham SE et al. (included UKB)*** |  |  |  |  |  |  |  |
| **Total cholesterol, SD** |  |  |  |  |  |  |  |
| Partially adjusted association^c^ | 805 | 0.178 (0.113, 0.243) | 1.13×10^-7^ | 1733 | 0.220 (0.177, 0.262) | 1.34×10^-23^ | 0.266 |
| Fully adjusted association^d^ | 641 | 0.197 (0.131, 0.264) | 9.17×10^-9^ | 1326 | 0.245 (0.199, 0.292) | 2.39×10^-24^ | 0.188 |
| **LDL cholesterol, SD** |  |  |  |  |  |  |  |
| Partially adjusted association^c^ | 805 | 0.210 (0.145, 0.275) | 5.21×10^-10^ | 1731 | 0.205 (0.163, 0.248) | 6.66×10^-21^ | 0.734 |
| Fully adjusted association^d^ | 641 | 0.216 (0.149, 0.283) | 4.58×10^-10^ | 1324 | 0.227 (0.181, 0.273) | 2.65×10^-21^ | 0.976 |
| **HDL cholesterol, SD** |  |  |  |  |  |  |  |
| Partially adjusted association^c^ | 742 | 0.285 (0.214, 0.356) | 1.33×10^-14^ | 1573 | 0.248 (0.208, 0.289) | 5.28×10^-32^ | 0.373 |
| Fully adjusted association^d^ | 586 | 0.322 (0.247, 0.397) | 3.96×10^-16^ | 1206 | 0.245 (0.204, 0.286) | 5.56×10^-30^ | 0.096 |
| **Triglycerides, SD** |  |  |  |  |  |  |  |
| Partially adjusted association^c^ | 805 | 0.288 (0.213, 0.362) | 1.15×10^-13^ | 1732 | 0.260 (0.211, 0.309) | 1.09×10^-24^ | 0.531 |
| Fully adjusted association^d^ | 641 | 0.291 (0.210, 0.371) | 4.40×10^-12^ | 1325 | 0.247 (0.193, 0.301) | 9.90×10^-19^ | 0.337 |
| ***PolyPred+*** |  |  |  |  |  |  |  |
| **Total cholesterol, SD** |  |  |  |  |  |  |  |
| Partially adjusted association^c^ | 805 | 0.259 (0.193, 0.324) | 2.46×10^-14^ | 1733 | 0.261 (0.219, 0.303) | 9.19×10^-33^ | 0.933 |
| Fully adjusted association^d^ | 641 | 0.278 (0.213, 0.344) | 5.02×10^-16^ | 1326 | 0.284 (0.238, 0.330) | 4.16×10^-32^ | 0.690 |
| **LDL cholesterol, SD** |  |  |  |  |  |  |  |
| Partially adjusted association^c^ | 805 | 0.273 (0.207, 0.338) | 1.29×10^-15^ | 1731 | 0.245 (0.202, 0.287) | 8.09×10^-29^ | 0.382 |
| Fully adjusted association^d^ | 641 | 0.290 (0.224, 0.355) | 3.60×10^-17^ | 1324 | 0.265 (0.220, 0.311) | 1.25×10^-28^ | 0.630 |
| **HDL cholesterol, SD** |  |  |  |  |  |  |  |
| Partially adjusted association^c^ | 742 | 0.348 (0.278, 0.418) | 6.96×10^-21^ | 1573 | 0.300 (0.260, 0.340) | 2.59×10^-46^ | 0.365 |
| Fully adjusted association^d^ | 586 | 0.383 (0.306, 0.460) | 6.74×10^-21^ | 1206 | 0.276 (0.235, 0.318) | 4.74×10^-36^ | 0.043 |
| **Triglycerides, SD** |  |  |  |  |  |  |  |
| Partially adjusted association^c^ | 805 | 0.251 (0.176, 0.326) | 9.82×10^-11^ | 1732 | 0.300 (0.251, 0.349) | 2.87×10^-32^ | 0.414 |
| Fully adjusted association^d^ | 641 | 0.253 (0.173, 0.333) | 1.19×10^-9^ | 1325 | 0.278 (0.224, 0.332) | 4.25×10^-23^ | 0.788 |

^a^ UKB, UK Biobank; SD, standard deviation; LDL, low-density lipoprotein; HDL, high-density lipoprotein.

^b^ *P*-value was obtained from the interaction term between lipid PGS and fish oil supplementation. Models were adjusted for lipids PGS, fish oil supplementation, sex, age, age^2^, assessment centers, genotyping array, the top 20 genetic principal components, body mass index, Townsend deprivation index, smoking status, alcohol status, physical activity, and statin use.

^c^ Models were adjusted for sex, age, age^2^, assessment centers, genotyping array, and the top 20 genetic principal components.

^d^ Models were adjusted for sex, age, age^2^, assessment centers, genotyping array, the top 20 genetic principal components, body mass index, Townsend deprivation index, smoking status, alcohol status, physical activity, and statin use.

**Supplementary Table 6.** Effects of fish oil supplementation and PGS on observed lipid levels in UKB participants of diverse ancestries^a^

|  | ***β*** | **SE** | ***P*-value** |
| --- | --- | --- | --- |
| **EUR** |  |  |  |
| ***Graham SE et al. (excluded UKB)*** |  |  |  |
| *Total cholesterol, SD* |  |  |  |
| Partially adjusted association^b^ |  |  |  |
| PGS | 0.258 | 0.001 | <2.0×10^−16^ |
| Fish oil intake | 0.055 | 0.003 | 1.12×10^-64^ |
| Fully adjusted association^c^ |  |  |  |
| PGS | 0.310 | 0.001 | <2.0×10^−16^ |
| Fish oil intake | 0.041 | 0.003 | 6.32×10^-39^ |
| *LDL cholesterol, SD* |  |  |  |
| Partially adjusted association^b^ |  |  |  |
| PGS | 0.275 | 0.001 | <2.0×10^−16^ |
| Fish oil intake | 0.040 | 0.003 | 2.57×10^-34^ |
| Fully adjusted association^c^ |  |  |  |
| PGS | 0.329 | 0.001 | <2.0×10^−16^ |
| Fish oil intake | 0.035 | 0.003 | 5.70×10^-29^ |
| *HDL cholesterol, SD* |  |  |  |
| Partially adjusted association^b^ |  |  |  |
| PGS | 0.232 | 0.001 | <2.0×10^−16^ |
| Fish oil intake | 0.076 | 0.003 | 1.37×10^-127^ |
| Fully adjusted association^c^ |  |  |  |
| PGS | 0.230 | 0.001 | <2.0×10^−16^ |
| Fish oil intake | 0.025 | 0.003 | 2.29×10^-14^ |
| *Triglycerides, SD* |  |  |  |
| Partially adjusted association^b^ |  |  |  |
| PGS | 0.261 | 0.001 | <2.0×10^−16^ |
| Fish oil intake | -0.020 | 0.003 | 2.24×10^-10^ |
| Fully adjusted association^c^ |  |  |  |
| PGS | 0.262 | 0.002 | <2.0×10^−16^ |
| Fish oil intake | 0.018 | 0.003 | 1.33×10^-7^ |
| ***Graham SE et al. (included UKB)*** |  |  |  |
| *Total cholesterol, SD* |  |  |  |
| Partially adjusted association^b^ |  |  |  |
| PGS | 0.284 | 0.001 | <2.0×10^−16^ |
| Fish oil intake | 0.054 | 0.003 | 1.19×10^-64^ |
| Fully adjusted association^c^ |  |  |  |
| PGS | 0.338 | 0.001 | <2.0×10^−16^ |
| Fish oil intake | 0.041 | 0.003 | 2.79×10^-39^ |
| *LDL cholesterol, SD* |  |  |  |
| Partially adjusted association^b^ |  |  |  |
| PGS | 0.293 | 0.001 | <2.0×10^−16^ |
| Fish oil intake | 0.040 | 0.003 | 3.86×10^-35^ |
| Fully adjusted association^c^ |  |  |  |
| PGS | 0.350 | 0.001 | <2.0×10^−16^ |
| Fish oil intake | 0.036 | 0.003 | 7.01×10^-30^ |
| *HDL cholesterol, SD* |  |  |  |
| Partially adjusted association^b^ |  |  |  |
| PGS | 0.324 | 0.001 | <2.0×10^−16^ |
| Fish oil intake | 0.077 | 0.003 | 8.15×10^-141^ |
| Fully adjusted association^c^ |  |  |  |
| PGS | 0.314 | 0.001 | <2.0×10^−16^ |
| Fish oil intake | 0.027 | 0.003 | 2.16×10^-18^ |
| *Triglycerides, SD* |  |  |  |
| Partially adjusted association^b^ |  |  |  |
| PGS | 0.284 | 0.001 | <2.0×10^−16^ |
| Fish oil intake | -0.020 | 0.003 | 3.10×10^-10^ |
| Fully adjusted association^c^ |  |  |  |
| PGS | 0.284 | 0.002 | <2.0×10^−16^ |
| Fish oil intake | 0.018 | 0.003 | 9.08×10^-8^ |
| ***Willer CJ et al.*** |  |  |  |
| *Total cholesterol, SD* |  |  |  |
| Partially adjusted association^b^ |  |  |  |
| PGS | 0.247 | 0.001 | <2.0×10^−16^ |
| Fish oil intake | 0.055 | 0.003 | 5.52×10^-64^ |
| Fully adjusted association^c^ |  |  |  |
| PGS | 0.295 | 0.001 | <2.0×10^−16^ |
| Fish oil intake | 0.041 | 0.003 | 1.25×10^-37^ |
| *LDL cholesterol, SD* |  |  |  |
| Partially adjusted association^b^ |  |  |  |
| PGS | 0.255 | 0.002 | <2.0×10^−16^ |
| Fish oil intake | 0.041 | 0.003 | 8.08×10^-36^ |
| Fully adjusted association^c^ |  |  |  |
| PGS | 0.304 | 0.001 | <2.0×10^−16^ |
| Fish oil intake | 0.036 | 0.003 | 5.89×10^-29^ |
| *HDL cholesterol, SD* |  |  |  |
| Partially adjusted association^b^ |  |  |  |
| PGS | 0.259 | 0.001 | <2.0×10^−16^ |
| Fish oil intake | 0.076 | 0.003 | 1.23×10^-129^ |
| Fully adjusted association^c^ |  |  |  |
| PGS | 0.256 | 0.001 | <2.0×10^−16^ |
| Fish oil intake | 0.025 | 0.003 | 2.93×10^-15^ |
| *Triglycerides, SD* |  |  |  |
| Partially adjusted association^b^ |  |  |  |
| PGS | 0.206 | 0.001 | <2.0×10^−16^ |
| Fish oil intake | -0.021 | 0.003 | 1.90×10^-10^ |
| Fully adjusted association^c^ |  |  |  |
| PGS | 0.208 | 0.002 | <2.0×10^−16^ |
| Fish oil intake | 0.018 | 0.003 | 2.05×10^-7^ |
| ***PolyPred*** |  |  |  |
| *Total cholesterol, SD* |  |  |  |
| Partially adjusted association^b^ |  |  |  |
| PGS | 0.416 | 0.001 | <2.0×10^−16^ |
| Fish oil intake | 0.050 | 0.003 | 2.15×10^-61^ |
| Fully adjusted association^c^ |  |  |  |
| PGS | 0.452 | 0.001 | <2.0×10^−16^ |
| Fish oil intake | 0.037 | 0.003 | 6.40×10^-38^ |
| *LDL cholesterol, SD* |  |  |  |
| Partially adjusted association^b^ |  |  |  |
| PGS | 0.421 | 0.001 | <2.0×10^−16^ |
| Fish oil intake | 0.037 | 0.003 | 1.94×10^-33^ |
| Fully adjusted association^c^ |  |  |  |
| PGS | 0.464 | 0.001 | <2.0×10^−16^ |
| Fish oil intake | 0.032 | 0.003 | 1.99×10^-28^ |
| *HDL cholesterol, SD* |  |  |  |
| Partially adjusted association^b^ |  |  |  |
| PGS | 0.533 | 0.001 | <2.0×10^−16^ |
| Fish oil intake | 0.062 | 0.003 | 1.85×10^-123^ |
| Fully adjusted association^c^ |  |  |  |
| PGS | 0.497 | 0.001 | <2.0×10^−16^ |
| Fish oil intake | 0.022 | 0.003 | 9.87×10^-16^ |
| *Triglycerides, SD* |  |  |  |
| Partially adjusted association^b^ |  |  |  |
| PGS | 0.449 | 0.001 | <2.0×10^−16^ |
| Fish oil intake | -0.016 | 0.003 | 1.17×10^-7^ |
| Fully adjusted association^c^ |  |  |  |
| PGS | 0.435 | 0.001 | <2.0×10^−16^ |
| Fish oil intake | 0.018 | 0.003 | 7.46×10^-9^ |
| ***P+T*** |  |  |  |
| *Total cholesterol, SD* |  |  |  |
| Partially adjusted association^b^ |  |  |  |
| PGS | 0.257 | 0.001 | <2.0×10^−16^ |
| Fish oil intake | 0.054 | 0.003 | 2.28×10^-63^ |
| Fully adjusted association^c^ |  |  |  |
| PGS | 0.301 | 0.001 | <2.0×10^−16^ |
| Fish oil intake | 0.040 | 0.003 | 8.51×10^-37^ |
| *LDL cholesterol, SD* |  |  |  |
| Partially adjusted association^b^ |  |  |  |
| PGS | 0.268 | 0.001 | <2.0×10^−16^ |
| Fish oil intake | 0.040 | 0.003 | 1.17×10^-34^ |
| Fully adjusted association^c^ |  |  |  |
| PGS | 0.314 | 0.001 | <2.0×10^−16^ |
| Fish oil intake | 0.035 | 0.003 | 1.26×10^-28^ |
| *HDL cholesterol, SD* |  |  |  |
| Partially adjusted association^b^ |  |  |  |
| PGS | 0.296 | 0.001 | <2.0×10^−16^ |
| Fish oil intake | 0.077 | 0.003 | 1.36×10^-136^ |
| Fully adjusted association^c^ |  |  |  |
| PGS | 0.291 | 0.001 | <2.0×10^−16^ |
| Fish oil intake | 0.026 | 0.003 | 4.74×10^-17^ |
| *Triglycerides, SD* |  |  |  |
| Partially adjusted association^b^ |  |  |  |
| PGS | 0.258 | 0.001 | <2.0×10^−16^ |
| Fish oil intake | -0.019 | 0.003 | 1.80×10^-9^ |
| Fully adjusted association^c^ |  |  |  |
| PGS | 0.259 | 0.002 | <2.0×10^−16^ |
| Fish oil intake | 0.019 | 0.003 | 1.71×10^-8^ |
| **AFR** |  |  |  |
| ***Graham SE et al. (excluded UKB)*** |  |  |  |
| *Total cholesterol, SD* |  |  |  |
| Partially adjusted association^b^ |  |  |  |
| PGS | 0.313 | 0.012 | 1.15×10^-145^ |
| Fish oil intake | 0.042 | 0.024 | 0.082 |
| Fully adjusted association^c^ |  |  |  |
| PGS | 0.352 | 0.013 | 1.44×10^-143^ |
| Fish oil intake | 0.011 | 0.027 | 0.675 |
| *LDL cholesterol, SD* |  |  |  |
| Partially adjusted association^b^ |  |  |  |
| PGS | 0.340 | 0.012 | 1.64×10^-167^ |
| Fish oil intake | 0.021 | 0.024 | 0.394 |
| Fully adjusted association^c^ |  |  |  |
| PGS | 0.385 | 0.013 | 1.25×10^-167^ |
| Fish oil intake | 0.007 | 0.027 | 0.789 |
| *HDL cholesterol, SD* |  |  |  |
| Partially adjusted association^b^ |  |  |  |
| PGS | 0.243 | 0.012 | 2.81×10^-91^ |
| Fish oil intake | 0.092 | 0.025 | 2.18×10^-4^ |
| Fully adjusted association^c^ |  |  |  |
| PGS | 0.233 | 0.013 | 6.33×10^-73^ |
| Fish oil intake | 0.056 | 0.027 | 0.040 |
| *Triglycerides, SD* |  |  |  |
| Partially adjusted association^b^ |  |  |  |
| PGS | 0.119 | 0.008 | 1.56×10^-46^ |
| Fish oil intake | -0.020 | 0.017 | 0.259 |
| Fully adjusted association^c^ |  |  |  |
| PGS | 0.129 | 0.010 | 1.72×10^-40^ |
| Fish oil intake | -0.012 | 0.021 | 0.566 |
| ***Graham SE et al. (included UKB)*** |  |  |  |
| *Total cholesterol, SD* |  |  |  |
| Partially adjusted association^b^ |  |  |  |
| PGS | 0.334 | 0.012 | 1.75×10^-163^ |
| Fish oil intake | 0.042 | 0.024 | 0.081 |
| Fully adjusted association^c^ |  |  |  |
| PGS | 0.375 | 0.013 | 2.90×10^-160^ |
| Fish oil intake | 0.011 | 0.027 | 0.690 |
| *LDL cholesterol, SD* |  |  |  |
| Partially adjusted association^b^ |  |  |  |
| PGS | 0.353 | 0.012 | 7.70×10^-178^ |
| Fish oil intake | 0.022 | 0.024 | 0.377 |
| Fully adjusted association^c^ |  |  |  |
| PGS | 0.399 | 0.013 | 1.52×10^-178^ |
| Fish oil intake | 0.007 | 0.027 | 0.792 |
| *HDL cholesterol, SD* |  |  |  |
| Partially adjusted association^b^ |  |  |  |
| PGS | 0.261 | 0.012 | 3.74×10^-106^ |
| Fish oil intake | 0.092 | 0.025 | 1.97×10^-4^ |
| Fully adjusted association^c^ |  |  |  |
| PGS | 0.250 | 0.013 | 1.61×10^-84^ |
| Fish oil intake | 0.056 | 0.027 | 0.038 |
| *Triglycerides, SD* |  |  |  |
| Partially adjusted association^b^ |  |  |  |
| PGS | 0.133 | 0.008 | 3.15×10^-58^ |
| Fish oil intake | -0.021 | 0.017 | 0.231 |
| Fully adjusted association^c^ |  |  |  |
| PGS | 0.140 | 0.010 | 3.96×10^-47^ |
| Fish oil intake | -0.013 | 0.020 | 0.537 |
| ***PolyPred+*** |  |  |  |
| *Total cholesterol, SD* |  |  |  |
| Partially adjusted association^b^ |  |  |  |
| PGS | 0.265 | 0.012 | 6.77×10^-110^ |
| Fish oil intake | 0.025 | 0.025 | 0.308 |
| Fully adjusted association^c^ |  |  |  |
| PGS | 0.288 | 0.013 | 9.63×10^-103^ |
| Fish oil intake | -0.007 | 0.028 | 0.790 |
| *LDL cholesterol, SD* |  |  |  |
| Partially adjusted association^b^ |  |  |  |
| PGS | 0.312 | 0.012 | 2.54×10^-149^ |
| Fish oil intake | 0.007 | 0.025 | 0.770 |
| Fully adjusted association^c^ |  |  |  |
| PGS | 0.338 | 0.013 | 8.12×10^-141^ |
| Fish oil intake | -0.012 | 0.028 | 0.658 |
| *HDL cholesterol, SD* |  |  |  |
| Partially adjusted association^b^ |  |  |  |
| PGS | 0.248 | 0.012 | 7.31×10^-97^ |
| Fish oil intake | 0.086 | 0.025 | 0.001 |
| Fully adjusted association^c^ |  |  |  |
| PGS | 0.242 | 0.013 | 7.19×10^-79^ |
| Fish oil intake | 0.053 | 0.027 | 0.050 |
| *Triglycerides, SD* |  |  |  |
| Partially adjusted association^b^ |  |  |  |
| PGS | 0.103 | 0.008 | 8.37×10^-35^ |
| Fish oil intake | -0.020 | 0.018 | 0.254 |
| Fully adjusted association^c^ |  |  |  |
| PGS | 0.104 | 0.010 | 2.51×10^-26^ |
| Fish oil intake | -0.009 | 0.021 | 0.674 |
| **CSA** |  |  |  |
| ***Graham SE et al. (excluded UKB)*** |  |  |  |
| *Total cholesterol, SD* |  |  |  |
| Partially adjusted association^b^ |  |  |  |
| PGS | 0.157 | 0.011 | 2.56×10^-49^ |
| Fish oil intake | 0.055 | 0.025 | 0.030 |
| Fully adjusted association^c^ |  |  |  |
| PGS | 0.207 | 0.011 | 4.68×10^-77^ |
| Fish oil intake | 0.071 | 0.026 | 0.006 |
| *LDL cholesterol, SD* |  |  |  |
| Partially adjusted association^b^ |  |  |  |
| PGS | 0.154 | 0.011 | 7.88×10^-48^ |
| Fish oil intake | 0.021 | 0.025 | 0.402 |
| Fully adjusted association^c^ |  |  |  |
| PGS | 0.203 | 0.011 | 7.84×10^-76^ |
| Fish oil intake | 0.051 | 0.026 | 0.048 |
| *HDL cholesterol, SD* |  |  |  |
| Partially adjusted association^b^ |  |  |  |
| PGS | 0.192 | 0.009 | 2.27×10^-108^ |
| Fish oil intake | 0.105 | 0.021 | 3.73×10^-7^ |
| Fully adjusted association^c^ |  |  |  |
| PGS | 0.191 | 0.010 | 8.59×10^-87^ |
| Fish oil intake | 0.056 | 0.023 | 0.013 |
| *Triglycerides, SD* |  |  |  |
| Partially adjusted association^b^ |  |  |  |
| PGS | 0.275 | 0.012 | 1.44×10^-114^ |
| Fish oil intake | -0.004 | 0.029 | 0.884 |
| Fully adjusted association^c^ |  |  |  |
| PGS | 0.264 | 0.014 | 4.27×10^-80^ |
| Fish oil intake | 0.032 | 0.032 | 0.332 |
| ***Graham SE et al. (included UKB)*** |  |  |  |
| *Total cholesterol, SD* |  |  |  |
| Partially adjusted association^b^ |  |  |  |
| PGS | 0.163 | 0.011 | 3.72×10^-53^ |
| Fish oil intake | 0.054 | 0.025 | 0.032 |
| Fully adjusted association^c^ |  |  |  |
| PGS | 0.215 | 0.011 | 8.82×10^-83^ |
| Fish oil intake | 0.071 | 0.026 | 0.006 |
| *LDL cholesterol, SD* |  |  |  |
| Partially adjusted association^b^ |  |  |  |
| PGS | 0.163 | 0.011 | 1.63×10^-53^ |
| Fish oil intake | 0.021 | 0.025 | 0.411 |
| Fully adjusted association^c^ |  |  |  |
| PGS | 0.215 | 0.011 | 3.57×10^-85^ |
| Fish oil intake | 0.052 | 0.026 | 0.041 |
| *HDL cholesterol, SD* |  |  |  |
| Partially adjusted association^b^ |  |  |  |
| PGS | 0.207 | 0.009 | 1.48×10^-125^ |
| Fish oil intake | 0.104 | 0.021 | 4.45×10^-7^ |
| Fully adjusted association^c^ |  |  |  |
| PGS | 0.209 | 0.009 | 1.44×10^-104^ |
| Fish oil intake | 0.053 | 0.022 | 0.018 |
| *Triglycerides, SD* |  |  |  |
| Partially adjusted association^b^ |  |  |  |
| PGS | 0.285 | 0.012 | 2.06×10^-123^ |
| Fish oil intake | -0.003 | 0.029 | 0.907 |
| Fully adjusted association^c^ |  |  |  |
| PGS | 0.276 | 0.014 | 2.68×10^-87^ |
| Fish oil intake | 0.032 | 0.032 | 0.321 |
| ***PolyPred+*** |  |  |  |
| *Total cholesterol, SD* |  |  |  |
| Partially adjusted association^b^ |  |  |  |
| PGS | 0.205 | 0.010 | 7.81×10^-85^ |
| Fish oil intake | 0.056 | 0.025 | 0.025 |
| Fully adjusted association^c^ |  |  |  |
| PGS | 0.278 | 0.011 | 8.27×10^-142^ |
| Fish oil intake | 0.074 | 0.025 | 0.003 |
| *LDL cholesterol, SD* |  |  |  |
| Partially adjusted association^b^ |  |  |  |
| PGS | 0.207 | 0.010 | 1.40×10^-85^ |
| Fish oil intake | 0.022 | 0.025 | 0.373 |
| Fully adjusted association^c^ |  |  |  |
| PGS | 0.283 | 0.011 | 3.07×10^-148^ |
| Fish oil intake | 0.056 | 0.025 | 0.026 |
| *HDL cholesterol, SD* |  |  |  |
| Partially adjusted association^b^ |  |  |  |
| PGS | 0.294 | 0.008 | 1.48×10^-262^ |
| Fish oil intake | 0.105 | 0.020 | 1.08×10^-7^ |
| Fully adjusted association^c^ |  |  |  |
| PGS | 0.300 | 0.009 | 2.09×10^-222^ |
| Fish oil intake | 0.053 | 0.021 | 0.013 |
| *Triglycerides, SD* |  |  |  |
| Partially adjusted association^b^ |  |  |  |
| PGS | 0.282 | 0.012 | 1.13×10^-120^ |
| Fish oil intake | -0.003 | 0.029 | 0.915 |
| Fully adjusted association^c^ |  |  |  |
| PGS | 0.271 | 0.014 | 2.46×10^-84^ |
| Fish oil intake | 0.030 | 0.032 | 0.349 |
| **EAS** |  |  |  |
| ***Graham SE et al. (included UKB)*** |  |  |  |
| *Total cholesterol, SD* |  |  |  |
| Partially adjusted association^b^ |  |  |  |
| PGS | 0.207 | 0.018 | 4.13×10^-30^ |
| Fish oil intake | 0.162 | 0.038 | 2.39×10^-5^ |
| Fully adjusted association^c^ |  |  |  |
| PGS | 0.231 | 0.019 | 1.11×10^-32^ |
| Fish oil intake | 0.138 | 0.040 | 0.001 |
| *LDL cholesterol, SD* |  |  |  |
| Partially adjusted association^b^ |  |  |  |
| PGS | 0.209 | 0.018 | 3.21×10^-30^ |
| Fish oil intake | 0.148 | 0.039 | 1.33×10^-4^ |
| Fully adjusted association^c^ |  |  |  |
| PGS | 0.227 | 0.019 | 2.37×10^-31^ |
| Fish oil intake | 0.130 | 0.040 | 0.001 |
| *HDL cholesterol, SD* |  |  |  |
| Partially adjusted association^b^ |  |  |  |
| PGS | 0.263 | 0.018 | 2.27×10^-46^ |
| Fish oil intake | 0.147 | 0.039 | 1.55×10^-4^ |
| Fully adjusted association^c^ |  |  |  |
| PGS | 0.269 | 0.018 | 2.13×10^-45^ |
| Fish oil intake | 0.096 | 0.040 | 0.018 |
| *Triglycerides, SD* |  |  |  |
| Partially adjusted association^b^ |  |  |  |
| PGS | 0.269 | 0.021 | 4.21×10^-37^ |
| Fish oil intake | 0.017 | 0.045 | 0.699 |
| Fully adjusted association^c^ |  |  |  |
| PGS | 0.263 | 0.023 | 3.34×10^-30^ |
| Fish oil intake | 0.067 | 0.049 | 0.167 |
| ***PolyPred+*** |  |  |  |
| *Total cholesterol, SD* |  |  |  |
| Partially adjusted association^b^ |  |  |  |
| PGS | 0.261 | 0.018 | 3.56×10^-46^ |
| Fish oil intake | 0.179 | 0.038 | 2.38×10^-6^ |
| Fully adjusted association^c^ |  |  |  |
| PGS | 0.286 | 0.019 | 3.37×10^-49^ |
| Fish oil intake | 0.159 | 0.039 | 5.49×10^-5^ |
| *LDL cholesterol, SD* |  |  |  |
| Partially adjusted association^b^ |  |  |  |
| PGS | 0.255 | 0.018 | 3.88×10^-44^ |
| Fish oil intake | 0.171 | 0.038 | 7.62×10^-6^ |
| Fully adjusted association^c^ |  |  |  |
| PGS | 0.277 | 0.019 | 4.57×10^-47^ |
| Fish oil intake | 0.156 | 0.040 | 8.13×10^-5^ |
| *HDL cholesterol, SD* |  |  |  |
| Partially adjusted association^b^ |  |  |  |
| PGS | 0.315 | 0.018 | 3.01×10^-66^ |
| Fish oil intake | 0.145 | 0.038 | 1.39×10^-4^ |
| Fully adjusted association^c^ |  |  |  |
| PGS | 0.301 | 0.019 | 2.03×10^-54^ |
| Fish oil intake | 0.092 | 0.040 | 0.022 |
| *Triglycerides, SD* |  |  |  |
| Partially adjusted association^b^ |  |  |  |
| PGS | 0.283 | 0.021 | 3.36×10^-41^ |
| Fish oil intake | 0.014 | 0.045 | 0.753 |
| Fully adjusted association^c^ |  |  |  |
| PGS | 0.267 | 0.023 | 2.85×10^-31^ |
| Fish oil intake | 0.064 | 0.049 | 0.187 |

^a^ UKB, UK Biobank; SD, standard deviation; LDL, low-density lipoprotein; HDL, high-density lipoprotein; EUR, European; AFR, African; CSA, Central/South Asian; EAS, East Asian.

^b^ Models were adjusted for sex, age, age^2^, assessment centers, genotyping array, and the top 20 genetic principal components.

^c^ Models were adjusted for sex, age, age^2^, assessment centers, genotyping array, the top 20 genetic principal components, body mass index, Townsend deprivation index, smoking status, alcohol status, physical activity, and statin use.

**Supplementary Figure 1**

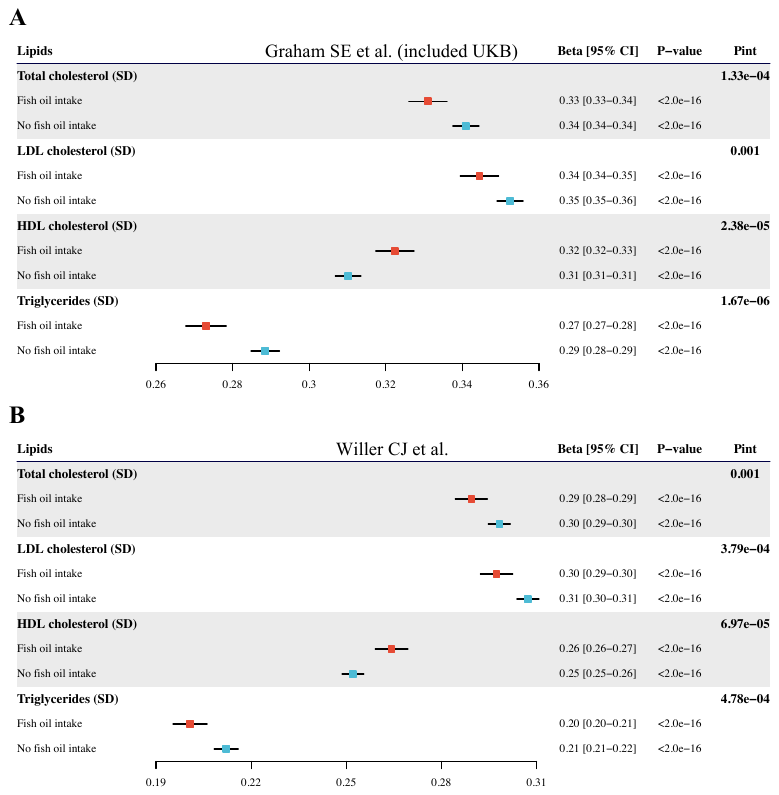

**Supplementary Figure 2**

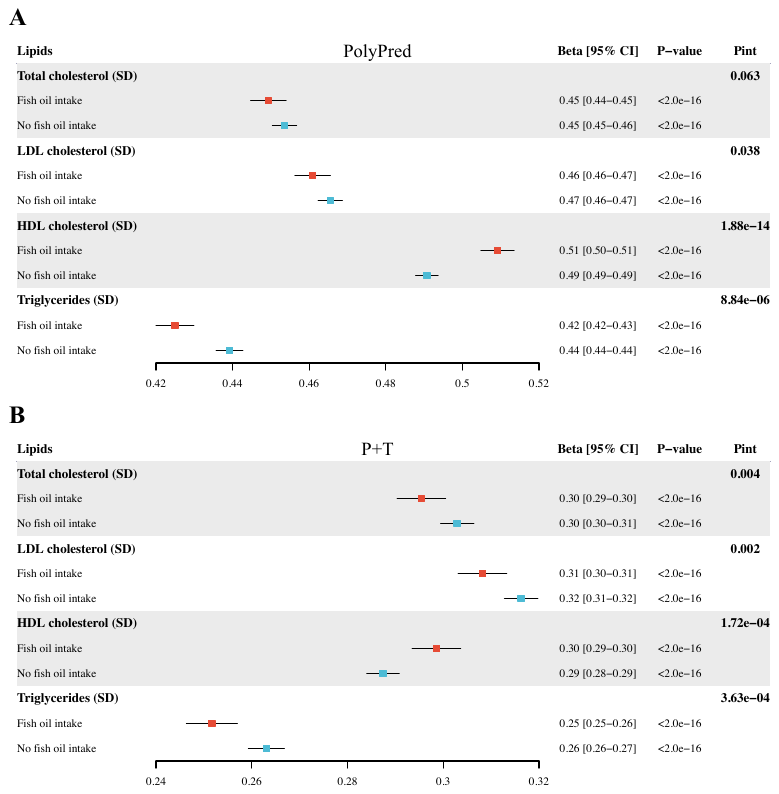
